# Detection without calibration: benchmarking domestic and international large language models for quality control of Mandarin ^18^F-FDG PET/CT reports

**DOI:** 10.64898/2026.06.24.26356406

**Authors:** Jingbo Wang, Weiqing Tang, Xingdi Ma, Huimin Yan, Ying Yuan

## Abstract

Large language models (LLMs) are increasingly used for automated quality control (QC) of radiology reports. However, the reliability of LLMs on reports in Mandarin, and the relative performance of domestic versus international flagship models, remain unknown. We benchmarked 14 LLM configurations, seven Chinese-developed (“domestic”) and seven international models, on 1,000 whole-body ^18^F-FDG PET/CT reports split into an error-injected “junior-doctor” arm and a low-residual “finalised” arm (500 each), using a controlled error-injection gold standard. Under each blinded zero-shot prompt, each model flagged six error types and assigned a 1–5 overall score. Two distinct abilities: error-detection macro-F1 (0.356–0.667) and overall-score calibration (ICC[2,1] 0.099–0.627), were weakly and not significantly correlated across models (Spearman ρ = 0.38, p = 0.18); the dissociation was instead evident in sharp rank reversals, the strongest detector (Claude-Opus-4.8 0.667) calibrating poorly (0.491), while the three best-calibrated models were all domestic (MiMo 0.627, GLM-5 0.612, DeepSeek 0.609). Once the access channel was controlled, domestic and international error detection were statistically indistinguishable (Δmacro-F1 = −0.011, P = 0.84); domestic models showed consistent but not significant advantages in calibration (ΔICC = +0.142) and Chinese-character-error detection (ΔF1 = +0.109), accompanied with large reductions in cost (US$0.09–2.71 vs $0.26–14.5 per 1,000 reports) and on-premise deployability. Re-running two flagships through both agent channels and clean APIs showed that agent channel inflated both detection and calibration (GPT-5.5 ΔICC = +0.098, 95% CI 0.070–0.128), confirming that uncontrolled benchmarks over-credit agent-channel models. Missed-diagnosis detection was the universal weakness (best 0.467) and the one category where the human physicians outperformed every model. Raw detection ability does not guarantee a trustworthy score, and domestic and international models differ by deployment-relevant profile rather than by overall performance rank; both essential distinctions for performing clinical nuclear-medicine QC.

## Introduction

Whole-body ^18^F-FDG PET/CT is central to the staging, restaging, treatment-response assessment and recurrence surveillance of malignancy. Unlike single-region examinations, a whole-body report must integrate anatomical and metabolic information across the entire body; it is longer, multi-layered and more information-dense with nuclear-medicine-specific terminology (glucose metabolism, uptake, standardized uptake value [SUV]). Report quality has a direct impact on clinical decision-making and patient safety, yet first-draft reports inevitably contain modality-inappropriate terminology, character-level typographical errors, left–right laterality contradictions and missed findings. Quality control (QC) today largely relies on line-by-line re-reading by senior physicians, a process that is inefficient, subjective and heavily drains already-scarce expert time.^1^

Large language models (LLMs) acquire robust natural-language processing from large-scale text pre-training and present as a promising solution to automate report QC.^2^ They have been applied to report generation, structured conversion, information extraction, quality control and patient communication,□,□,^1^□□^2^□,^23^□^2^□, and have reportedly reached near-expert-level performance on several clinical-text tasks.□ For radiology reports in English, GPT-4 detects report errors at comparable performance as radiologists while significantly shortening review time and cost.^3^ For reports in Mandarin, one study has compared multiple LLMs for QC of head-and-neck CT-angiography reports, with generally high error-detection F1 values□ and report classification, summarisation and generation by LLMs have also been explored.^1^□,^21,22^ Whole-body PET/CT reports are substantially longer than single-region reports, span more anatomical regions, and interleave metabolic information with nuclear-medicine terminology, thus demand far more advanced cross-segmental semantic understanding. However, LLM-based QC of nuclear-medicine PET/CT reports in Mandarin remains largely understudied. To our knowledge, no study has benchmarked LLM error-detection QC on whole-body PET/CT reports; the nearest QC benchmarks address other modalities such as chest radiographs and CT reports.^31^

In addition to the lack of LLM application in whole-body report QC, two additional considerations motivate this study. First, deploying LLM in report QC brings two distinct but complementing functions: error detection, where LLMs flag specific defects, and score calibration, which pertains to quality ratings that track ground truth. A model that flags defects well may still score reports unreliably, however, these two abilities are rarely evaluated and reported together. Clinically they serve different ends: error flags tell a reviewer what to fix, whereas a calibrated score is what a downstream triage system acts on, so strength on one need not imply the other. Currently, hospitals conduct report QC mainly by relying on senior radiologists to review a few randomly sampled reports, which serve as an approximation of the entire reports collection. If LLMs could be reliably implemented, hospitals could conduct report QC on all reports in a cost-efficient manner instead of lengthy, labour-intensive manual review of only a few reports. Second, the *deployability* of an LLM in a regulated clinical workflow depends on factors orthogonal to raw capability, for instance, language fidelity to clinical text in Mandarin, inference cost and latency, and the governance of patient data and access to the model itself etc. These considerations bear unevenly on domestically developed (Chinese) versus international flagship models: domestic models are trained on a larger proportion of Chinese text and can often be deployed on-premise, whereas international flagships usually lead in general-purpose benchmarks, but are proprietary, costlier, and often subject to access-policy changes beyond the control of individual institutions, which might lead to risks in data safety, operational dependencies and management. While prior work has shown that privacy-compliant, open-source models already approach proprietary commercial models in radiology error detection^12^, no study has yet compared domestic and international LLMs head-to-head on Mandarin clinical-text QC, nor examined whether detection and calibration capabilities diverge to advise clinics in decision making regarding LLM deployment.

Here we benchmark a balanced roster of LLMs (seven domestic and seven international configurations) for QC of Mandarin whole-body ^18^F-FDG PET/CT reports. After using controlled error injection to establish a known gold standard over two report arms (an error-injected “junior-doctor” arm and a low-residual “finalised” arm), each model was deployed under identical blinded zero-shot prompts, to flag six error types and assign a five-point overall quality score. We (i) quantified per-category and macro-averaged error-detection F1 and overall-score calibration (intraclass correlation, ICC), with bootstrap confidence intervals; (ii) tested whether detection and calibration are dissociated across models; (iii) characterized the performance of domestic versus international models across detection, calibration, Chinese-character-error sensitivity, over-flagging, cost, latency and deployment governance; and (iv) anchored all models against a human expert baseline read by four attending nuclear-medicine physicians (two with 10 years’ and two with 5 years’ of experience). Our central finding is that error detection and score calibration are distinct capabilities, and that domestic and international models differ less by overall rank than by practical considerations relevant to real-world clinical implementation. Rather than crowning a single best model, this study provides a reference point for where current large language models stand on quality control of Mandarin whole-body PET/CT reports.

## Results

### Overall error detection

We evaluated fourteen LLM configurations (seven Chinese-developed and seven international) on the identical 1,000 reports, split into two 500-report arms (Fig. 1); full specifications, versions, access channels and pricing are given in Methods and Supplementary Appendix S1. To prevent access channel from confounding the comparison, the Anthropic and OpenAI models were run both through their original agent channels and through the same clean API used for the other models, with the two agent-channel configurations reported separately throughout. Bootstrap 95% confidence intervals (2,000 arm-stratified resamples) accompany the headline metrics.

**Fig. 1.**
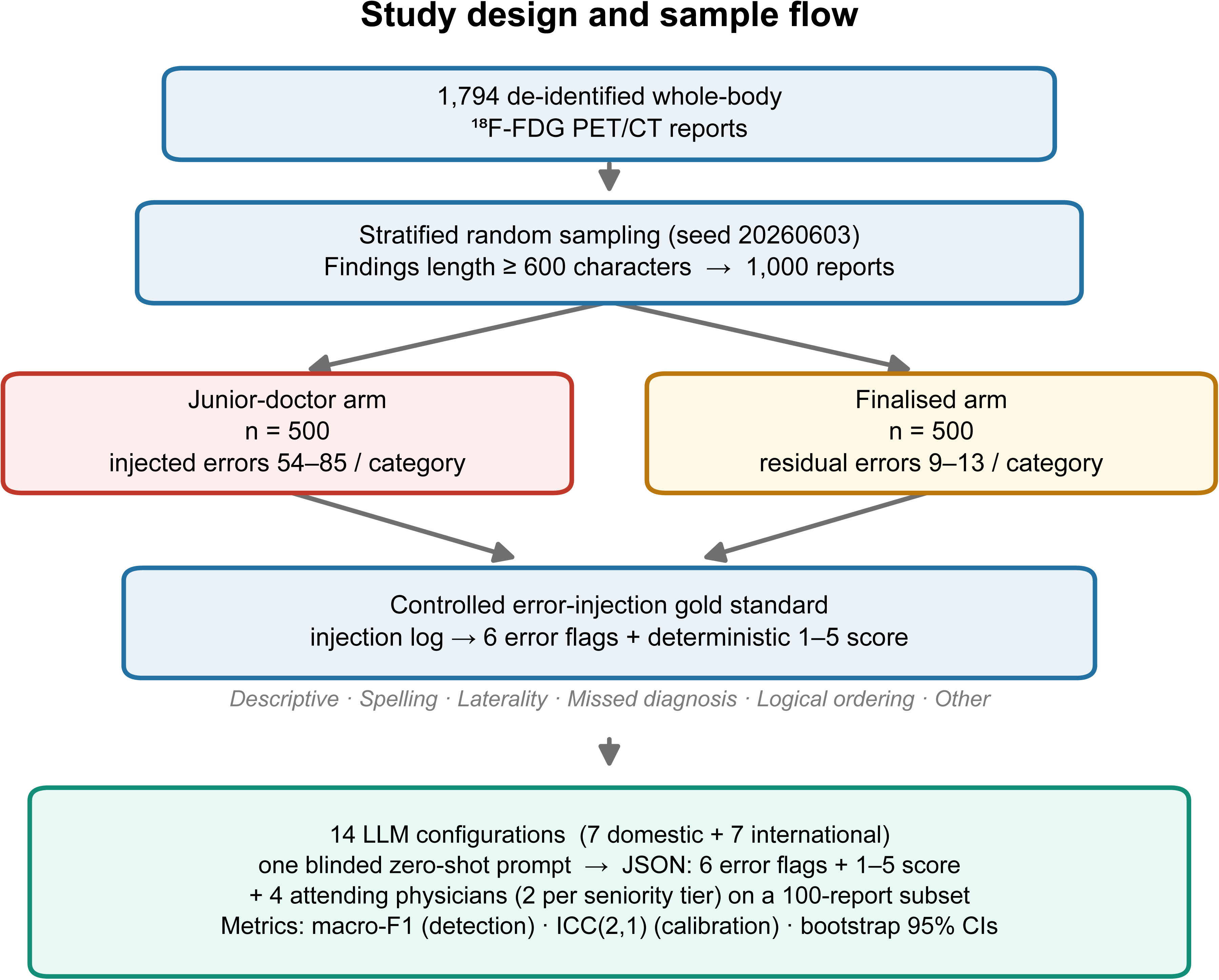
Study design and sample flow. 1,794 de-identified whole-body ^18^F-FDG PET/CT reports were filtered (Findings ≥ 600 characters) and stratified-randomly sampled (seed 20260603) to 1,000 reports, split into a 500-report junior-doctor arm (injected errors) and a 500-report finalised arm (residual errors). Controlled error injection yields the gold standard (six error flags + a deterministic 1–5 score). Fourteen LLM configurations (seven domestic + seven international) and four attending nuclear-medicine physicians (two per seniority tier, on a 100-report subset) were evaluated under one identical blinded zero-shot prompt.

Macro-averaged F1 (mean of six per-category F1 values, pooled, n = 1,000) spanned 0.356 (Grok-4.3) to 0.667 (Claude-Opus-4.8) (Table 1). The clean-API Claude-Opus-4.8 was the single strongest detector, narrowly above the agent-channel Claude-Fable-5 (0.660); the leading domestic models (MiMo 0.602, DeepSeek 0.594, GLM-5 0.586) trailed the front-runners by 0.06–0.08. Critically, when the formal comparison was restricted to the 12 clean-API configurations (pre-specified; agent-channel models excluded), domestic and international detection were statistically indistinguishable (domestic median macro-F1 0.507 vs international 0.567; Δ = −0.011 [95% CI −0.024 to +0.001]; permutation P = 0.84): the international detection lead was carried by the two agent-channel flagships, and among clean-API international models the macro-F1 range (Grok 0.356 to Opus-4.8 0.667) fully overlapped the domestic range (see below).

**Table 1.**
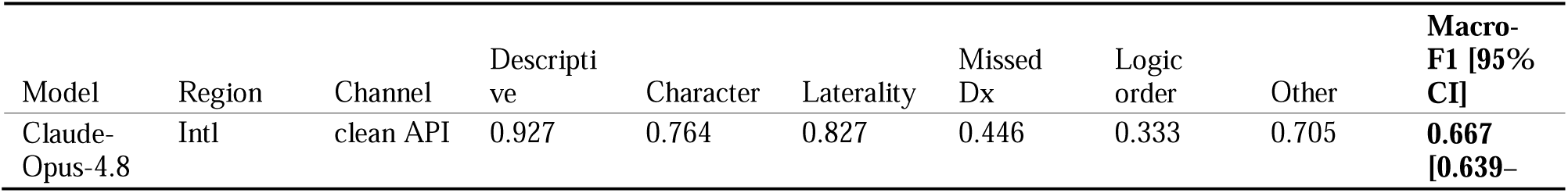
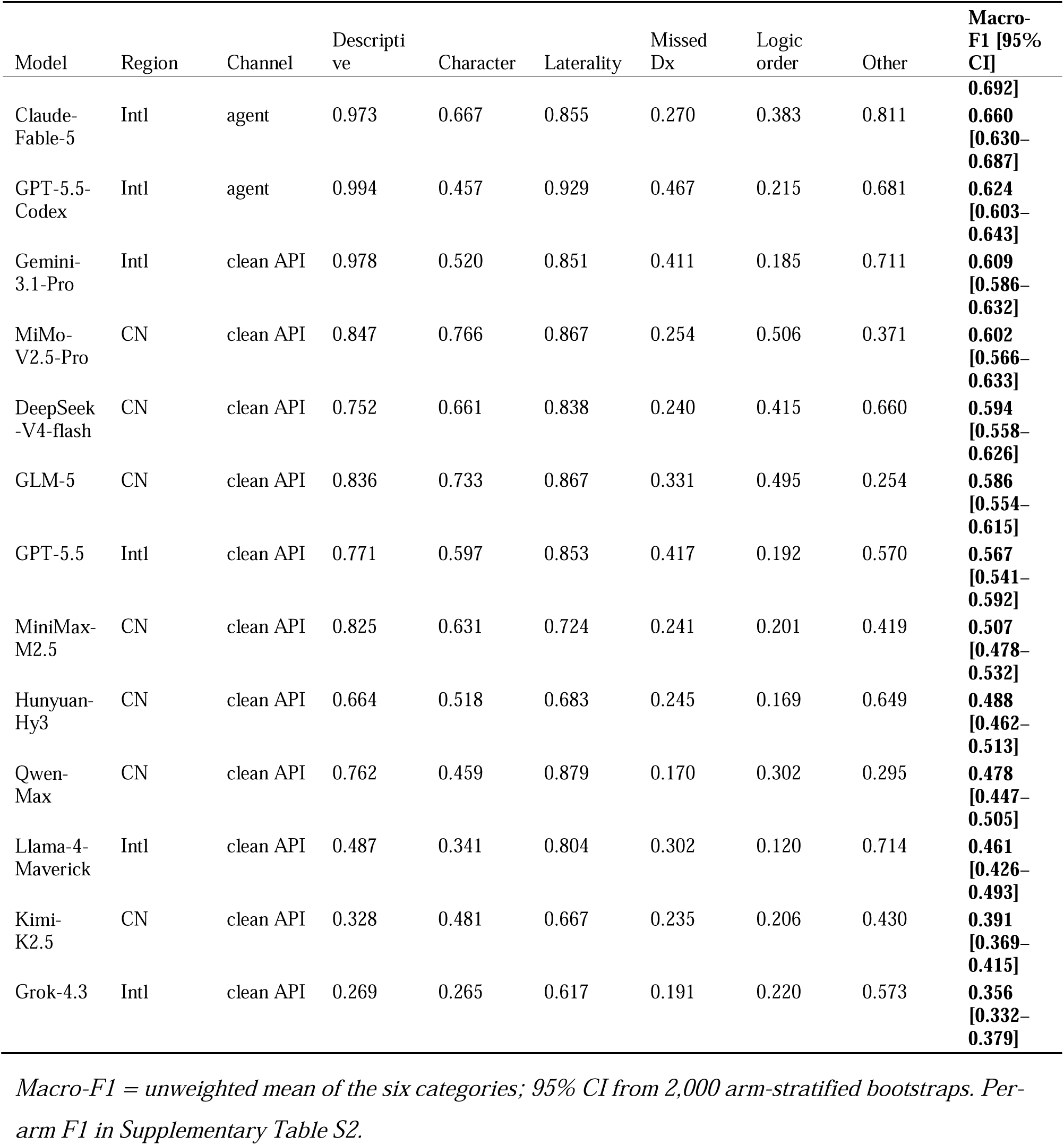
Per-category and macro-averaged error-detection F1 (pooled, n = 1,000), ranked by macro-F1.

### Detection–calibration distinction

Agreement between each model’s overall 1–5 score and the gold-standard score (ICC[2,1]; two-way random, single measure, absolute agreement) ranged from 0.099 to 0.627 (Table 2; all P < 0.001). On the Koo & Li (2016) scale, five models reached *moderate* agreement (0.50–0.75) and nine were *poor*; none reached *good* (≥ 0.75).

**Table 2.** Overall-score agreement with the gold standard, ICC(2,1), ranked by overall ICC.

| Model | Region | Channel | Junior arm | Finalized arm | Overall [95% CI] | Agreement |
| --- | --- | --- | --- | --- | --- | --- |
| MiMo-V2.5-Pro | CN | clean API | 0.564 | 0.547 | **0.627 [0.573–0.674]** | Moderate |
| GLM-5 | CN | clean API | 0.564 | 0.489 | **0.612 [0.562–0.657]** | Moderate |
| DeepSeek-V4-flash | CN | clean API | 0.596 | 0.447 | **0.609 [0.555–0.656]** | Moderate |
| Claude-Fable-5 | Intl | agent | 0.636 | 0.323 | **0.559 [0.513–0.607]** | Moderate |
| Llama-4-Maverick | Intl | clean API | 0.484 | 0.456 | **0.544 [0.484–0.592]** | Moderate |
| Claude-Opus-4.8 | Intl | clean API | 0.561 | 0.255 | **0.491 [0.446–0.536]** | Poor |
| MiniMax-M2.5 | CN | clean API | 0.479 | 0.209 | **0.414 [0.359–0.469]** | Poor |
| Hunyuan-Hy3 | CN | clean API | 0.458 | 0.165 | **0.369 [0.322–0.415]** | Poor |
| Qwen-Max | CN | clean API | 0.367 | 0.210 | **0.352 [0.306–0.394]** | Poor |
| GPT-5.5-Codex | Intl | agent | 0.398 | 0.141 | **0.329 [0.294–0.364]** | Poor |
| Kimi-K2.5 | CN | clean API | 0.342 | 0.109 | **0.262 [0.224–0.302]** | Poor |
| Gemini-3.1-Pro | Intl | clean API | 0.337 | 0.079 | **0.244 [0.212–0.276]** | Poor |
| GPT-5.5 | Intl | clean API | 0.301 | 0.093 | **0.231 [0.198–0.263]** | Poor |
| Grok-4.3 | Intl | clean API | 0.144 | 0.036 | **0.099 [0.078–0.119]** | Poor |

*All $P < 0.001$ (F-test). Note the systematic drop from junior to finalised arm (e.g. Claude-Fable-5 0.636 → 0.323), where low error prevalence and compressed score variance make calibration harder; MiMo degraded least (0.564 → 0.547). 95% CI from 2,000 bootstraps.*

Calibration was largely orthogonal to detection (Fig. 2a): across the 14 models macro-F1 and ICC were only weakly and non-significantly correlated (Spearman ρ = 0.38, p = 0.18). The three best-calibrated models were all domestic (MiMo 0.627, GLM-5 0.612, DeepSeek 0.609), while the international flagships tend to rank lower (GPT-5.5-Codex 0.329, Gemini 0.244, GPT-5.5-API 0.231, Grok 0.099). Models tend to perform well in only one of the two axes while poorly in the other: the strongest detector, Claude-Opus-4.8, fell to 6th on calibration (0.491), and Gemini from 4th in detection to 12th in calibration, whereas MiMo rose from 5th in detection to 1st in calibration. This single-axis competence is the crux for deployment: a usable QC tool must both find errors and score them reliably, so excelling on one axis alone is insufficient. Only Claude-Fable-5 (detection 2nd, calibration 4th) and MiMo (5th, 1st) ranked in the top five on both axes—one international, one domestic. The dissociation, and the regional calibration pattern, are quantified below (Domestic versus international models).

**Fig. 2.**
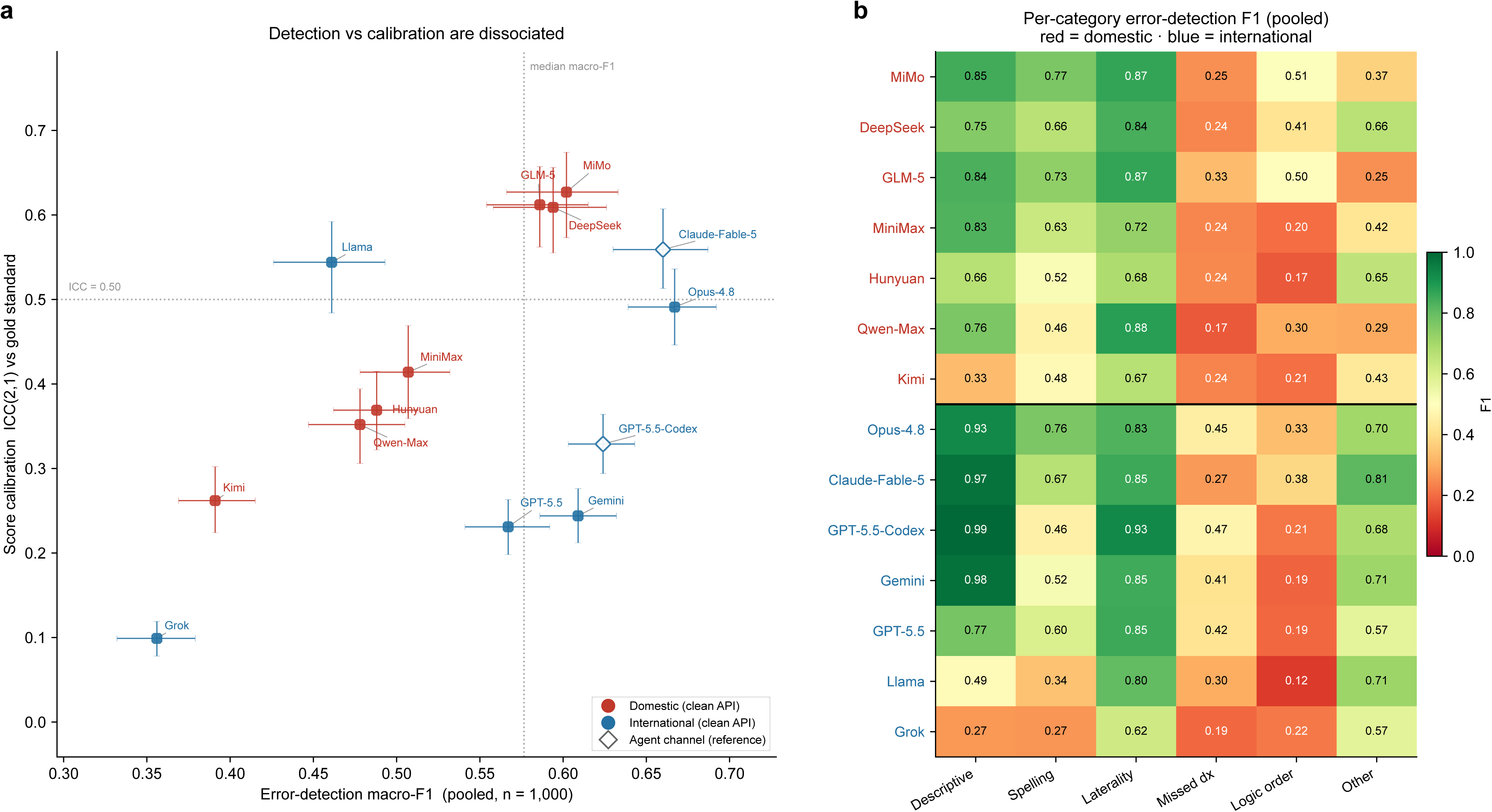
Detection and calibration across the 14 models. **(a)** Error-detection macro-F1 versus score-calibration ICC(2,1): one point per model, coloured by region (red = domestic, blue = international), with distinct markers for the open-weight model (filled square) and the agent-channel reference points (open diamonds), bootstrap 95% CI whiskers, and quadrant guides at the median macro-F1 and ICC = 0.50. Detection ability and calibration are dissociated. **(b)** Per-category error-detection F1 (pooled, n = 1,000) as a 14 × 6 heat-map; rows grouped by region (domestic above, international below the black divider) and ordered by macro-F1 within group.

### Error-type-specific patterns

Performance varied far more across error types than across models (Table 1, Fig. 2b). Descriptive (terminology) and laterality errors were structurally simple and the most detectable for most models (0.994 and 0.929, GPT-5.5-Codex; Claude-Opus-4.8 0.927 and 0.827). Only Grok (0.269) and Kimi (0.328) failed on descriptive errors, both through gross over-flagging (see below). Chinese-character-error detection is championed by domestic models, though comparable performance can be achieved by the latest international model: the top three were domestic (MiMo 0.766, GLM-5 0.733, DeepSeek 0.661), but Claude-Opus-4.8 scored similarly (0.764), while the other international models scored lower (Gemini 0.520, GPT-5.5 0.597/0.457). Detection of homophone/visually-similar character errors thus favours models with strong Mandarin exposure and training, regardless of model origin. Missed-diagnosis detection was the universal weakness for all models: the best score was only 0.467 (GPT-5.5-Codex; Claude-Opus-4.8 0.446, GPT-5.5-API 0.417), and most scores fell below 0.31. Because missed diagnoses are the most clinically consequential errors, this is the one category where humans excel and thus the most crucial limitation of LLM-driven QC automation. And lastly, logical-ordering detection was uniformly weak, with low precision across the board (best MiMo 0.506, GLM-5 0.495; ten models below 0.34), driven by pervasive over-flagging (see below).

### Over-flagging and score severity explain the dissociation

The calibration failures documented above were driven by over-reporting of errors and overly harsh scoring (table1_error_counts.csv, table3_score_dist.csv). Several models flagged errors that far exceeded that of the gold standard: Qwen-Max flagged missed diagnosis in 394 (junior) and 413 (finalised) reports versus gold counts of 75 and 12, reporting an omission in nearly every report, which explains its missed-diagnosis F1 of 0.170; Grok over-flagged across the board; and logical ordering was over-flagged by most models. The same models scored too harshly: on the finalised arm (86.8% gold-score 5), Grok gave a 2 to 59.4% of reports, and GPT-5.5 (both channels), Gemini and Claude-Opus-4.8 all scored a mode of 3 (clean-API GPT-5.5 71.4% at score 3, Opus-4.8 44.2% at 3). By contrast the best-calibrated domestic models tracked the gold distribution (finalised arm, MiMo 85.6% at 5 vs gold 86.8%; GLM-5 71.0%, DeepSeek 70.0%). Models that both over-flag and score severely produce overall scores that no longer track the reference—precisely why their ICC collapsed (Fig. 3a).

**Fig. 3.**
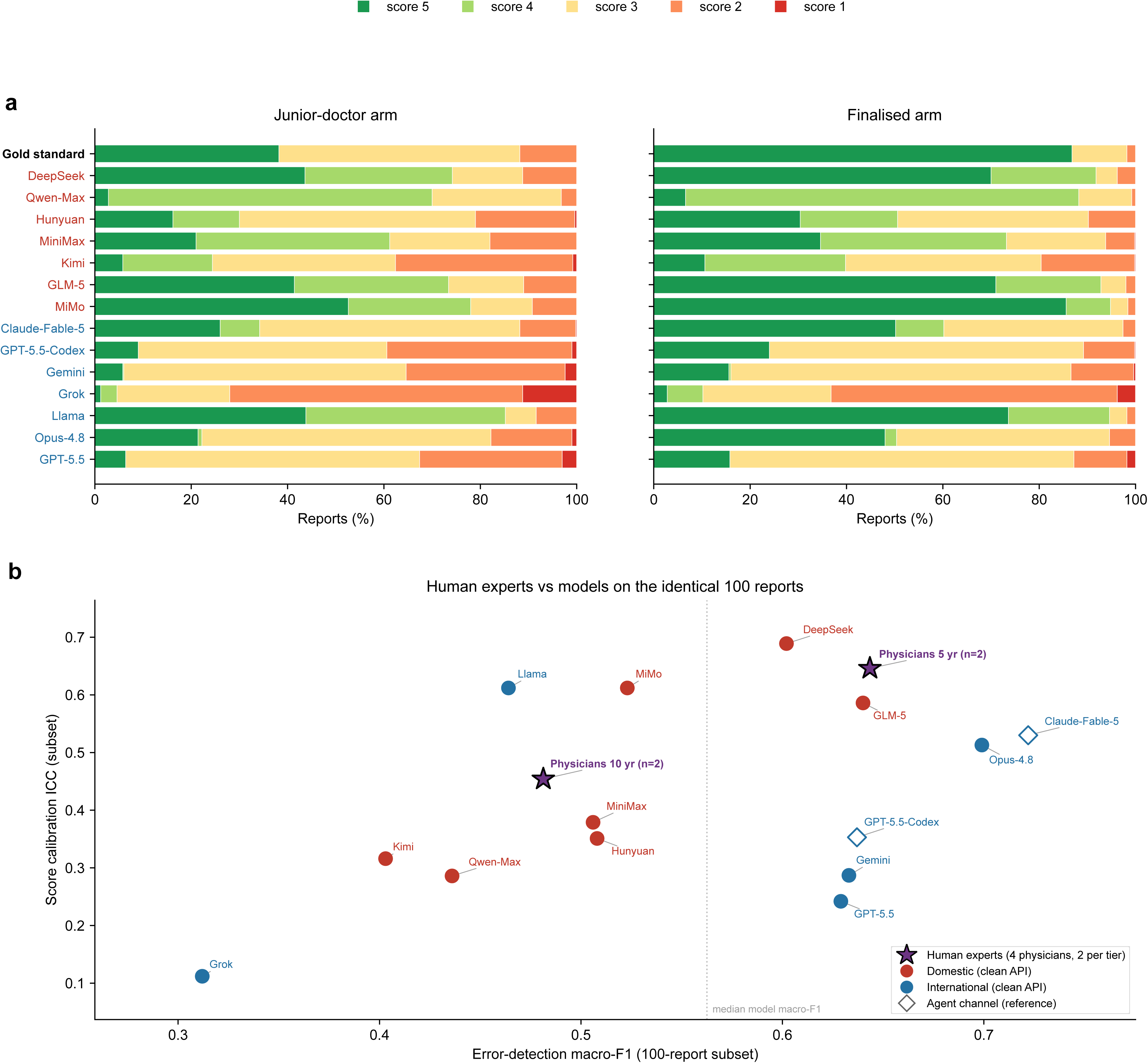
Scoring behaviour and the human baseline. **(a)** Distribution of the overall 1–5 score versus the gold standard for every model in each arm (junior-doctor and finalised); the gold-standard row is in bold and model rows are coloured by region (red = domestic, blue = international); the score key (1 = red to 5 = green) is shared by both arms. **(b)** Human experts versus models on the identical 100-report subset: error-detection macro-F1 versus calibration ICC for all 14 models and the four attending physicians. Marker scheme (key in panel): purple stars = human experts (pooled 10-year and 5-year groups, n = 2 each); filled circles = clean-API models coloured by region; filled square = open-weight Llama; open diamonds = agent-channel models (Claude-Fable-5, GPT-5.5-Codex); the dotted vertical line marks the median model macro-F1.

### Efficiency and cost

We summarised input/output tokens, per-report latency and cost per 1,000 reports for the twelve clean-API configurations (Table 3; efficiency_summary.csv). The two agent-channel configurations (Claude-Fable-5, GPT-5.5-Codex) produce no comparable logs and are omitted—but their clean-API counterparts (Claude-Opus-4.8, GPT-5.5) now supply Anthropic and OpenAI cost/latency directly, while earlier agent-only access could not.

**Table 3.**
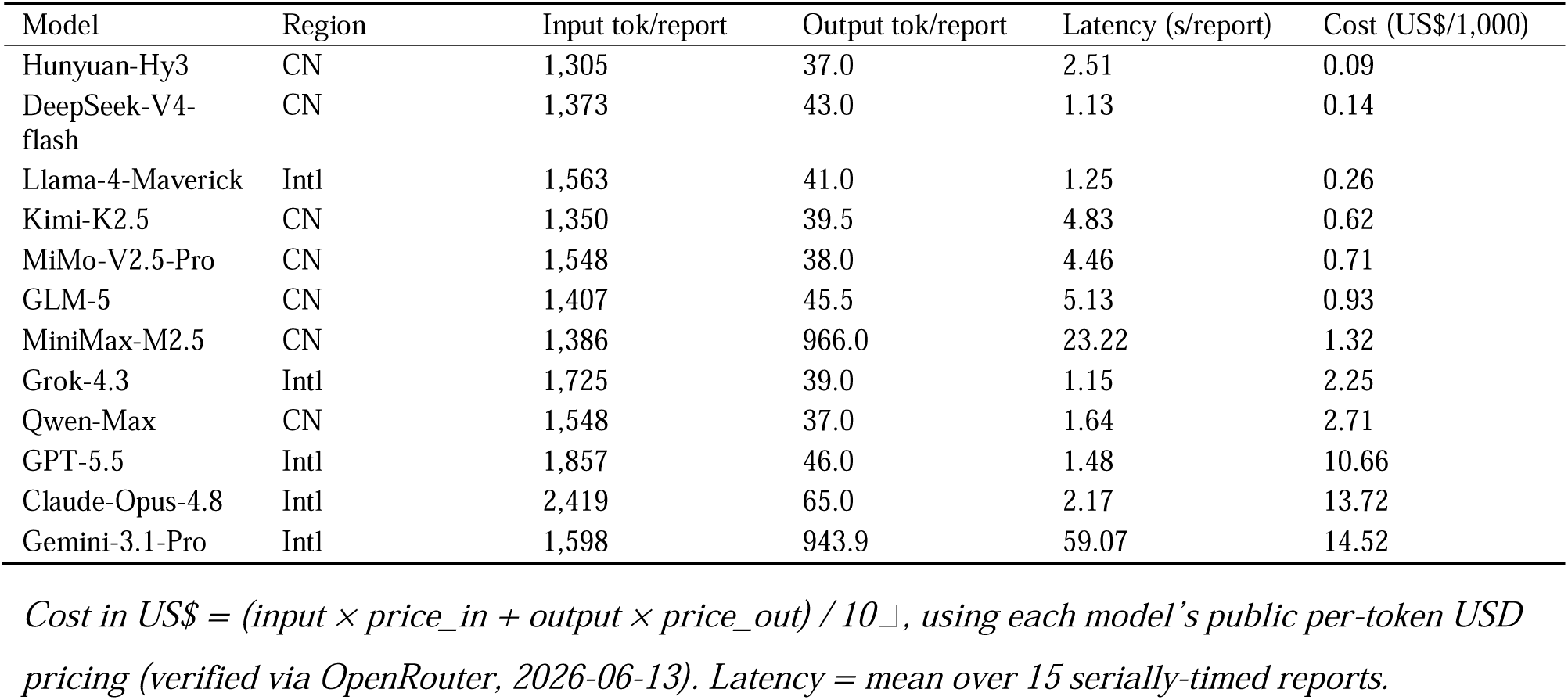
Efficiency and cost for clean-API configurations (per 1,000 reports), ranked by cost.

Per-1,000-report cost varied more than 150-fold, from US$0.09 to $14.52, which is governed by per-token price, output verbosity and tokeniser efficiency rather than model origin. The cheapest were domestic (Hunyuan-Hy3 $0.09, DeepSeek-V4-flash $0.14) and the open international Llama ($0.26); the costliest were the international flagships (Gemini $14.52, Claude-Opus-4.8 $13.72, GPT-5.5 $10.66), inflated both by forced reasoning (Gemini ∼944 output tokens/report) and by less efficient tokenization of Mandarin—the clean-API international models consumed 1.4–1.8× more input tokens than domestic models for identical prompts (e.g. Claude-Opus-4.8 2,419 vs DeepSeek 1,373 tokens/report). Latency tracked reasoning mode, not region: the reasoning-forced Gemini (59.1 s/report) and MiniMax (23.2 s) were slowest, while DeepSeek (1.13 s), Grok (1.15 s), Llama (1.25 s), GPT-5.5 (1.48 s) and Claude-Opus-4.8 (2.17 s) were all 1–2 s.

### Human expert baseline and human–model comparison

Four attending nuclear-medicine physicians, two with 10 years’ and two with 5 years’ PET/CT reporting experience, independently applied the identical QC instrument to a stratified random subset of 100 reports (60 injected-error, 40 finalised), blinded to injection status. Within each seniority pair, one physician rated the first 50 reports of the subset and the other the remaining 50 (each report thus receiving one 10-year and one 5-year rating); ratings were pooled per seniority group for analysis. All 14 models were re-scored on these same 100 reports; subset values (Table 5) differ from the full-cohort results and are not cross-read with them.

A careful human expert is among the best-balanced readers. The 5-year group reached macro-F1 0.643 and ICC 0.646, only slightly behind the two Claude configurations on detection and behind DeepSeek (0.689) on calibration—and both seniority groups outperformed every model on missed-diagnosis detection (5-year group 0.759, 10-year group 0.583 vs best model 0.556), the models’ universal weakness. The 10-year and 5-year experience physician readings agreed only poorly (between-physician ICC 0.44), yet the best models’ calibration (subset ICC up to 0.689) already meets or exceeds this human ceiling, suggesting that report-quality scoring is intrinsically subjective. The senior (10-year) pair did not outscore the junior (5-year) pair on this QC instrument; with only two physicians per tier, this is underpowered and likely reflects individual reading style (the 10-year pair flagged more conservatively higher precision, lower recall) rather than a expertise effect. Humans nonetheless retain a clear edge on clinically critical omissions, defining a complementary division of labour (Fig. 3b).

### Domestic versus international models: a profile difference, not a ranking

All 14 model configurations were evaluated on the identical 1,000 reports. The formal region test was pre-specified on the 12 clean-API configurations; the two agent-channel models (Claude-Fable-5, GPT-5.5-Codex) are shown only as reference points (Fig. 2a).

Model origin did not cleanly separate the models (Table 4, Fig. 2a). On the clean-API set, error detection did not differ between regions: domestic macro-F1 (median 0.507, range 0.391–0.602) and international macro-F1 (median 0.567, range 0.356–0.667) were statistically indistinguishable (Δ = −0.011 [95% CI−0.024 to +0.001]; permutation P = 0.84). The apparent international detection advantage seen in uncontrolled comparisons was carried by the two agent-channel flagships (channel-effect analysis below) and did not survive once access channel was controlled; among clean-API international models the macro-F1 range (Grok 0.356 to Claude-Opus-4.8 0.667) fully overlaps the domestic range.

**Table 4.**
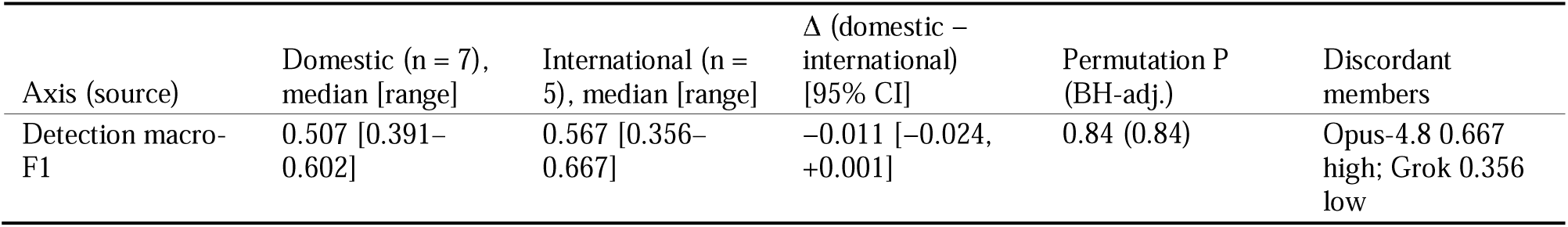
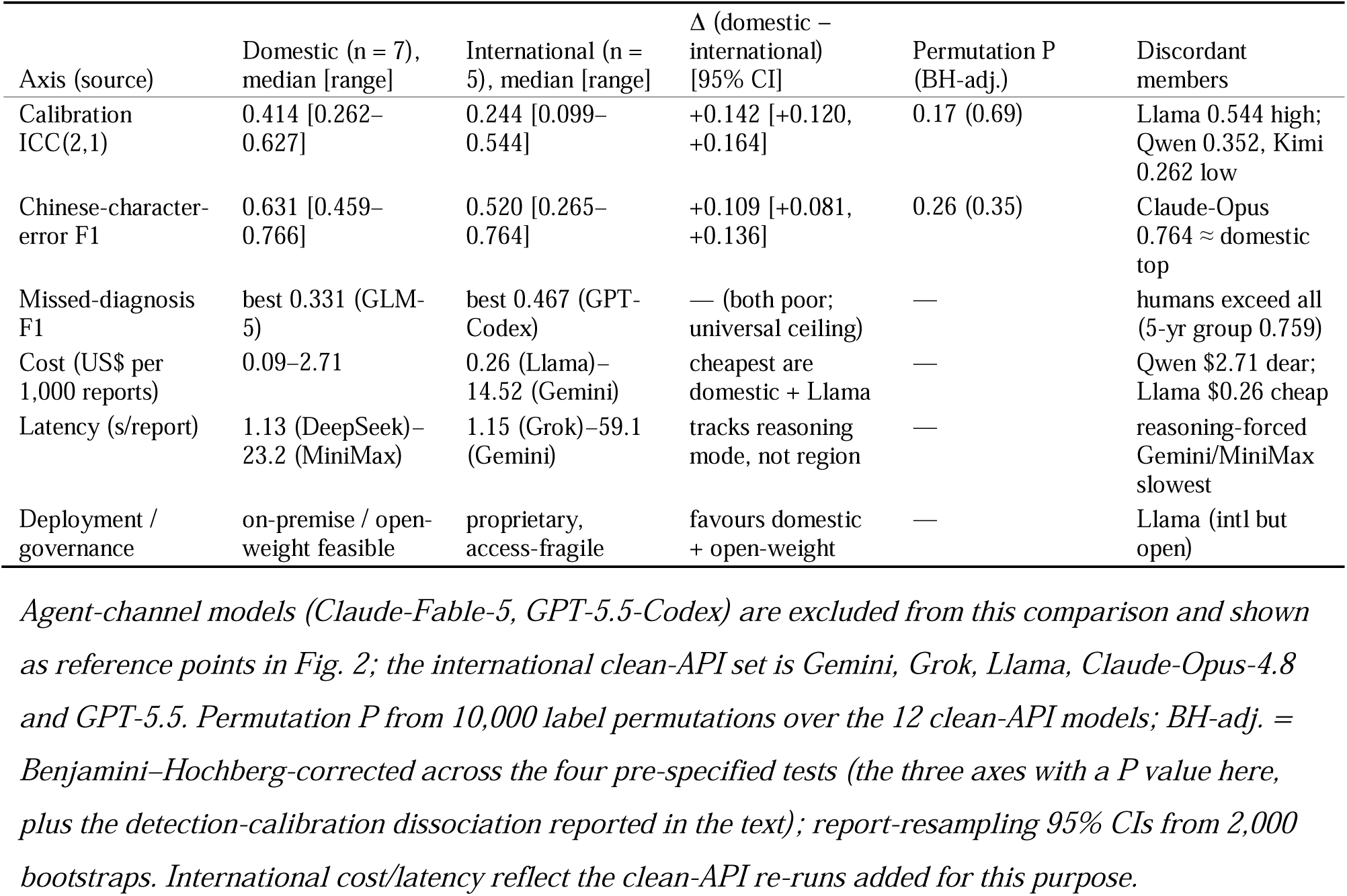
Domestic versus international models across axes (clean-API set; n = 1,000 reports).

**Table 5.**
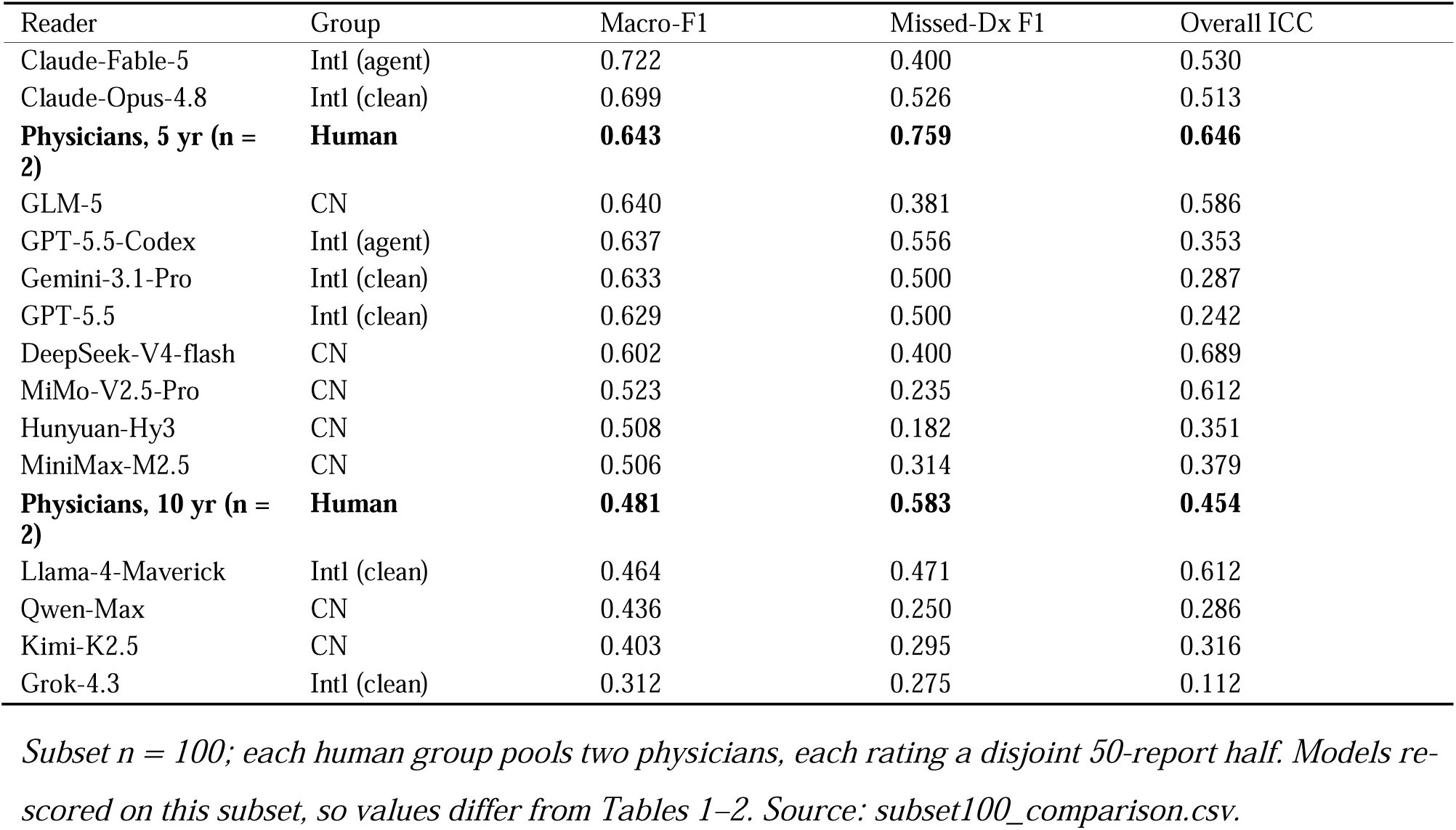
Human raters and all 14 models on the identical 100-report subset, ranked by macro-F1.

| Reader | Group | Macro-F1 | Missed-Dx F1 | Overall ICC |
| --- | --- | --- | --- | --- |
| Claude-Fable-5 | Intl (agent) | 0.722 | 0.400 | 0.530 |
| Claude-Opus-4.8 | Intl (clean) | 0.699 | 0.526 | 0.513 |
| **Physicians, 5 yr (n = 2)** | **Human** | **0.643** | **0.759** | **0.646** |
| GLM-5 | CN | 0.640 | 0.381 | 0.586 |
| GPT-5.5-Codex | Intl (agent) | 0.637 | 0.556 | 0.353 |
| Gemini-3.1-Pro | Intl (clean) | 0.633 | 0.500 | 0.287 |
| GPT-5.5 | Intl (clean) | 0.629 | 0.500 | 0.242 |
| DeepSeek-V4-flash | CN | 0.602 | 0.400 | 0.689 |
| MiMo-V2.5-Pro | CN | 0.523 | 0.235 | 0.612 |
| Hunyuan-Hy3 | CN | 0.508 | 0.182 | 0.351 |
| MiniMax-M2.5 | CN | 0.506 | 0.314 | 0.379 |
| **Physicians, 10 yr (n = 2)** | **Human** | **0.481** | **0.583** | **0.454** |
| Llama-4-Maverick | Intl (clean) | 0.464 | 0.471 | 0.612 |
| Qwen-Max | CN | 0.436 | 0.250 | 0.286 |
| Kimi-K2.5 | CN | 0.403 | 0.295 | 0.316 |
| Grok-4.3 | Intl (clean) | 0.312 | 0.275 | 0.112 |

Subset n = 100; each human group pools two physicians, each rating a disjoint 50-report half. Models re-scored on this subset, so values differ from Tables 1–2. Source: subset100\_comparison.csv.

Model origin influenced performance mainly in score calibration and Chinese-character-error detection, both of which favoured domestic models. Overall-score calibration was higher for domestic models (ICC median 0.414 vs 0.244; Δ = +0.142 [95% CI +0.120 to +0.164]; permutation P = 0.17), and the three best-calibrated models were all domestic (MiMo 0.627, GLM-5 0.612, DeepSeek 0.609). Detection of Chinese-character errors likewise favoured domestic models (F1 median 0.631 vs 0.520; Δ = +0.109 [95% CI +0.081 to +0.136]; permutation P = 0.26; top three all domestic, though Claude-Opus-4.8 matched them at 0.764). The report-resampling CIs for both differences excluded zero, but the model-level permutation tests did not reach P < 0.05 and did not survive Benjamini–Hochberg correction across the pre-specified tests (Table 4; adjusted P > 0.35) as between-model variance within each region is large. The open-weight international Llama calibrated well [ICC 0.544] while the international flagships Gemini, Grok and GPT-5.5 calibrated poorly [0.099–0.244]. Both findings are therefore consistent, sizeable but not formally significant regional tendencies, potentially reflecting the influence of greater Chinese-language and Chinese-clinical-text exposure during pre-training.

Model origin was not a clean predictor of performance, and four counterexamples make the point: Llama-4-Maverick is international yet open-weight, inexpensive and moderately calibrated; Claude-Opus-4.8 (international) matched the domestic best on Chinese-character-error detection and calibrated better than four of the seven domestic models; Qwen-Max and Kimi (domestic) were among the worst over-flaggers and poorest calibrators; and Grok-4.3 (international) ranked last on both axes. Our results suggest that key considerations of LLM performance, such as Mandarin familiarity, conservative/calibrated scoring, cost, and on-premise governability, correlate with but are not determined by model origin. The deployment-relevant distinction is well-calibrated, cheap, governable (often domestic or open-weight) versus proprietary, access-fragile, over-flagging tendencies (as seen in several international flagships), an axis that only partly aligns with domestic versus international.

These profile differences are sharpened by cost, latency and governance (Table 4; Methods). Per-1,000-report cost spanned more than 150-fold, and the cheapest models were all domestic (Hunyuan $0.09, DeepSeek $0.14) together with the open international model (Llama $0.26), whereas the international flagships were the costliest (Gemini ≈$14.5, Claude-Opus-4.8 ≈$13.7, GPT-5.5 ≈$10.7 per 1,000)—inflated both by per-token pricing and by less efficient tokenisation of Chinese (the clean-API international models consumed 1.4–1.8× more input tokens than domestic models for the identical prompts; e.g. Claude-Opus-4.8 2,419 vs DeepSeek 1,373 tokens/report). Latency, by contrast, tracked reasoning mode, not region: the reasoning-forced Gemini (59.1 s/report) and MiniMax (23.2 s) were by far the slowest, while DeepSeek (1.13 s), Grok (1.15 s), Llama (1.25 s), GPT-5.5 (1.48 s) and Claude-Opus-4.8 (2.17 s) were all 1–2 s. Finally, locally deployable models, namely domestic models and the open-weight Llama, can run on-premise, a prerequisite where PET/CT report text cannot leave the institution and where uninterrupted, auditable access is required; proprietary frontier models do not share this property.

### Channel-effect analysis

Because Claude-Fable-5 and GPT-5.5-Codex were reached through agent channels rather than the clean API, we re-ran the Anthropic and OpenAI models through the identical clean OpenRouter channel. For OpenAI this is a same-model, two-channel comparison and isolates the channel effect: relative to the clean API, the Codex agent channel inflated both GPT-5.5’s detection (macro-F1 0.624 vs 0.567; Δ = +0.057 [95% CI +0.038 to +0.076]) and its calibration (ICC 0.329 vs 0.231; Δ = +0.098 [95% CI +0.070 to +0.128]). The Anthropic comparison (Claude-Fable-5 agent vs Claude-Opus-4.8 clean API) confounds channel with model but points the same way for calibration (ICC 0.559 vs 0.491; Δ = +0.069 [95% CI +0.019 to +0.117]; detection essentially unchanged, Δ = −0.007). The access channel therefore materially affects measured performance, justifying its exclusion from the formal region test and confirming that the headline regional findings (no regional detection difference; calibration and character-error detection favouring domestic models) are robust to it.

## Discussion

In this study, we benchmarked 14 large language model configurations for quality control of Mandarin whole-body ^18^F-FDG PET/CT reports. Our results showed that the two abilities a QC model must possess—detecting specific errors and assigning a calibrated overall score—are dissociated, and that domestic and international models differ less by overall capability than by practical considerations relevant to clinical deployment. Across the roster, error-detection macro-F1 and score-calibration ICC were only weakly and non-significantly correlated (Spearman ρ = 0.38, p = 0.18), and the rank orderings reversed sharply: the strongest detector (Claude-Opus-4.8, macro-F1 0.667) calibrated poorly (ICC 0.491), whereas the three best-calibrated models were all domestic (MiMo 0.627, GLM-5 0.612, DeepSeek 0.609) despite mid-pack ranking in detection. Raw error-detection ability therefore does not guarantee a trustworthy overall score, because a single holistic score is what a downstream triage system would most plausibly act on.

The observed dissociation is most likely due to over-flagging coupled with overly harsh scoring. Several models reported errors far exceeding errors injected in the gold standard—most strikingly Qwen-Max, which flagged a missed diagnosis in 394/500 junior and 413/500 finalised reports against gold counts of 75 and 12—and then assigned correspondingly severe overall scores. On the finalised arm, where 86.8% of reports are error-free, the harshest models (Grok, Gemini, GPT-5.5 in both channels) graded the large majority as “2” or “3”. Over-flagging inflates recall but collapses precision, and severe scoring detaches the overall rating from the reference; the two together are precisely why calibration failed for the flagship brands. In a clinical QC setting, this false-positive burden is not benign—every spurious flag consumes the senior-physician review time that automation is meant to save.

Beyond these brand-level scoring failures, a consistent pattern emerged across models: performance varied far more by error type than by model. Errors decidable from local text features—laterality, modality-inappropriate terminology and Chinese-character errors—were detected well (F1 up to 0.93), whereas errors requiring cross-segment semantic alignment were not. Missed-diagnosis detection, which demands linking an abnormality described in the Findings to its absence from the Impression at the two ends of a long report, was the universal weakness: the best model reached only 0.467, and most fell below 0.31. This category is also the most clinically consequential, and it is the one where the human physicians retained a clear edge (5-year group 0.759), with both seniority groups exceeding every model. Unlike human misses, which arise from satisfaction-of-search and premature-closure biases□, the models’ failures reflect a limitation of long-range, discourse-level reasoning rather than perceptual oversight—consistent with the difficulty of the same category in single-region CT-angiography reports.□ A concurrent benchmark formalised this same divide between pattern-based errors, decidable from local linguistic form, and reasoning-dependent errors that require domain knowledge or long-range integration: in German-origin English CT, MRI and X-ray reports, GPT-4.1 and Llama-3.3-70B likewise failed most on physiologically impossible findings (two-stage success as low as 1.9%).³^2^ Our missed-diagnosis result extends this convergent gap to Mandarin whole-body PET/CT and a 14-model roster, locating it at the long-range, discourse-integration end of the reasoning-dependent spectrum, while adding the calibration axis and human baseline that the two-model study lacked.

While it is commonly assumed that international flagships would outperform domestic models, once the access channel was controlled their detection performance did not differ significantly. In the clean-API set, domestic and international macro-F1 were statistically indistinguishable (Δ = −0.011, permutation P = 0.84). The performance differed only in score calibration (domestic higher by ΔICC = 0.142) and Chinese-character-error detection (domestic higher by ΔF1 = 0.109). Differences in both factors reproduced under report-resampling (bootstrap CIs excluded zero), but did not survive multiple-comparison correction at the model level (Benjamini–Hochberg-adjusted P > 0.35), potentially due to the small number of models tested and large within-region variance. These observed differences are therefore consistent and sizeable but not statistically significant, plausibly reflecting greater exposure to Mandarin clinical text during pre-training in domestic models. Model origin was not deterministic: the open-weight international model Llama calibrated better than four domestic models; Claude-Opus-4.8 matched the domestic best on Chinese-character-error detection (0.764); and the domestic Qwen-Max and Kimi were among the worst over-flaggers. The deployment-relevant axis is thus a behavioural profile—calibrated, inexpensive, governable—that correlates with, but is not reducible to, national origin.

Cost is a decisive practical constraint for clinical deployment, and it varied enormously. Per-1,000-report cost spanned more than 150-fold across the 14 models: the cheapest were domestic (Hunyuan $0.09, DeepSeek $0.14) and open-weight Llama ($0.26), while the international flagships were the costliest (Gemini ≈$14.5, Claude-Opus-4.8 ≈$13.7, GPT-5.5 ≈$10.7 per 1,000). Two factors compounded the price: forced reasoning (Gemini, MiniMax) and, more notably, less efficient tokenisation of Mandarin, with the international clean-API models consuming 1.4–1.8× more input tokens than domestic models for identical prompts. Latency, by contrast, tracked reasoning mode rather than region: the reasoning-forced Gemini (59 s/report) and MiniMax (23 s) were the slowest, while DeepSeek, Grok, Llama, GPT-5.5 and Opus were all 1–2 s. Separately, because two flagships were originally reached through agent channels (Claude Code, ChatGPT Codex) rather than a clean API, we re-ran the Anthropic and OpenAI models through the identical clean channel, and performance shifted. For OpenAI, a same-model two-channel comparison, the agent channel inflated both detection (ΔF1 = 0.057, 95% CI 0.038–0.076) and calibration (ΔICC = 0.098, 95% CI 0.070–0.128) relative to the API. The Anthropic comparison (agent Fable-5 vs clean-API Opus-4.8) confounds channel with model but agrees for calibration (ΔICC = +0.069), with detection essentially unchanged. Access channel is therefore not a neutral implementation detail: in this one clean same-model comparison the agent channel over-credited the model on both axes, so the regional findings reported here are stated on the channel-controlled set.

For real-time, on-premise QC, where PET/CT report text cannot leave the institution and uninterrupted, auditable access is required, locally deployable models—domestic systems and open-weight Llama—have a structural advantage that the strongest proprietary detector cannot offset. Access to proprietary frontier models can also be curtailed by policy outside the deploying institution’s control; a marginally stronger model that cannot be reliably or compliantly accessed is of limited use in a regulated workflow. A pragmatic deployment is therefore sequential: a fast, cheap, well-calibrated model (often domestic or open-weight) is first deployed to screen surface errors, escalating only high-risk or ambiguous reports to a second, reasoning-focused model, with a final false-positive verification step—an approach shown to roughly double positive predictive value at lower cost.^1^□

Against the human baseline, the best models’ calibration was comparable to or better than the agreement between the physicians: the four physicians (two per seniority tier) agreed only poorly on the overall score (between-physician ICC 0.44), a ceiling that the best models already meet or exceed (subset ICC up to 0.689 for DeepSeek). Holistic report-quality scoring is intrinsically subjective, and apparent model “miscalibration” must be judged against a human ceiling that is itself only poor-to-moderate. At the same time, humans retained a clear advantage in detecting clinically critical omissions, defining a complementary division of labour rather than replacement. The more senior (10-year) pair did not outperform the junior (5-year) pair on this structured QC task—an underpowered, individual-style observation (n = 2 per tier) rather than a demonstrated seniority effect—reinforcing how subjective holistic report-quality scoring is.

This study has several limitations. While controlled error injection enables a known gold standard set, which is impossible to achieve with unaltered reports, the injected error distribution may differ from that of naturally occurring errors, therefore, external validation on real pre-review reports is needed. The data are single-centre and weighted toward head-and-neck tumour reports (a general tertiary hospital with a prominent head-and-neck oncology service; ∼94% of reports concern head-and-neck primaries, and the whole-body findings concentrate there), so generalisation to other diseases, regions and modalities requires multi-centre confirmation; the design’s strength is internal validity through a consistent gold standard, and the dissociation is a model-behaviour finding less tied to case-mix. Evaluation was zero-shot—the deployable baseline that isolates intrinsic ability; few-shot prompting and domain fine-tuning are known to raise error detection substantially^13^,^1^□ and define the improvement roadmap. Results are sensitive to prompt design and to the inherently subjective definition of “omission”. Channel non-equivalence was directly measured and the regional inferences restricted to the clean-API set, but residual heterogeneity remains (Gemini and MiniMax could not have reasoning disabled, and the current frontier Anthropic/OpenAI models do not accept a temperature parameter, so a temperature-0 setting identical to the other models is unattainable for them). Finally, the regional comparison is underpowered at the model level (7 vs 5), and the finalised-arm ICC is deflated by low score variance; both temper the strength of the regional claims. The human baseline comprises only four readers at a single centre under a disjoint-half design (each physician rated 50 of the 100 reports), so within-tier inter-rater reliability could not be computed and the human ceiling should not be over-generalised.

## Conclusion

For automated quality control of Mandarin whole-body ^18^F-FDG PET/CT reports, large language models detect surface and structured errors reasonably well but miss semantically deep errors, above all the clinically critical missed diagnosis. Most score reports with only poor-to-moderate calibration, detection ability and score calibration are dissociated. Domestic and international models differ by practical implementation considerations rather than overall rank in performance: once the access channel is controlled, the two groups detect errors equally well. Our results showed that domestic and open-weight models offer better score calibration and Chinese-character-error detection, far lower cost, on-premise governability, and access stability, against a marginally higher detection from the international flagships that comes with poorer calibration, higher cost and policy-dependent access that can be fragile and unstable. The pragmatic choice for clinical nuclear-medicine QC is a calibrated, inexpensive, locally deployable model (often domestic or open-weight) embedded in a human-in-the-loop cascade, with senior physicians retained for the omission detection that models have yet to match.

## Methods

### Report source and de-identification

This retrospective study was approved by the Institutional Review Board of Shanghai Ninth People’s Hospital, Shanghai Jiao Tong University School of Medicine (approval number SH9H-2026-T390-1), which waived informed consent because only de-identified text was used. Finalised whole-body ^18^F-FDG PET/CT reports were exported anonymously by the information department; reports with missing or insufficient imaging description or impression were excluded, yielding a working pool of 1,794 reports. On export, all direct identifiers—name, examination number, inpatient/outpatient number and reporting physician—were removed, and residual names or identity numbers in free text were scrubbed. Only the “Findings” (检查所见) and “Impression” (检查结论) sections were retained; no clinical history was included.

### Sampling and two-arm design

Reports shorter than 600 characters in the Findings section were filtered out to ensure sufficient content. From the remaining pool, 1,000 reports were drawn by stratified random sampling (fixed seed 20260603 for reproducibility) and split into two disjoint 500-report arms, mirroring the 500 + 500 design of prior work□: a junior-doctor arm simulating unreviewed first drafts, and a finalised arm simulating senior-physician reviewed reports.

### Error taxonomy and overall score

Following prior studies^3^,□ and the characteristics of PET/CT reports, six error types were defined: (1) descriptive errors: use of other-modality terminology for metabolic findings, e.g. “signal” (MRI) or “enhancement” in place of “glucose metabolism/uptake”; (2) character errors: Chinese-character typos, especially in organ/anatomical names (e.g. 甲装腺 for 甲状腺 [thyroid], 淋巴节 for 淋巴结 [lymph node], 纵膈 for 纵隔 [mediastinum]); (3) laterality errors: left/right contradictions within the report or between Findings and Impression; (4) missed-diagnosis errors: an abnormality clearly described in Findings but absent from the Impression; (5) logical-ordering errors: Impression items not following the conventional cranial-to-caudal anatomical sequence (brain → head and neck → chest → abdomen and pelvis → skeleton); and (6) other errors: unit errors (e.g. attaching a dimension to the unitless SUV, confusing cm with mm), punctuation and date errors. Descriptive, character, missed-diagnosis, logical-ordering and other errors were classified as *common* errors and laterality as a *special* error. Overall report quality was rated on a five-point Likert scale: 5 = no error, logically coherent; 4 = no error, slightly imperfect language; 3 = one or two common errors; 2 = three common errors or any special error; 1 = more than three common errors with a special error.

### Controlled error injection (gold standard)

Because finalised reports contain very few native errors, a controlled error-injection method was used to establish a known gold standard^3^. For the junior-doctor arm (n = 500), each of the six error types was injected programmatically into reports at a pre-set per-report probability (descriptive 0.15, character 0.15, laterality 0.12, missed-diagnosis 0.18, logical-ordering 0.15, other 0.15), at most one instance per type per report to permit a per-report × per-type binary formulation. The finalised arm (n = 500) received residual errors at one-fifth of these rates, simulating post-review reports. Every injection recorded the error type, the original text and the modification; this log constitutes the gold standard, from which a deterministic reference Likert score was derived by the rubric above. The resulting injected-positive counts (junior / finalised) were: descriptive 76 / 13, character 77 / 13, laterality 54 / 9, missed-diagnosis 75 / 12, logical-ordering 61 / 11, other 85 / 13.

### Models and access channels

To avoid subjective selection, domestic models were drawn from an objective third-party usage ranking: as of June 2026, the public leaderboard of the LLM-aggregation platform OpenRouter (openrouter.ai/rankings, ranked by platform-wide tokens processed) showed Chinese-developed models accounting for over half of platform traffic, with the seven selected models consistently near the top. We therefore included seven domestic models—DeepSeek-V4-flash, Qwen-Max (Alibaba), Hunyuan-Hy3 (Tencent), MiniMax-M2.5, Kimi-K2.5 (Moonshot AI), GLM-5 (Zhipu AI) and MiMo-V2.5-Pro (Xiaomi)—and seven international model configurations comprising the four leading closed-source flagships (Anthropic, OpenAI, Google, xAI) plus an open-weights representative (Meta Llama-4-Maverick).

Models were reached through three access channels, disclosed in full for transparency (Supplementary Table S1): (i) a clean OpenAI-compatible API—vendor-direct for the three domestic models with first-party endpoints (DeepSeek, Qwen-Max, Hunyuan) and via OpenRouter for the other four domestic models plus Gemini-3.1-Pro, Grok-4.3 and Llama-4-Maverick—each at temperature 0 with one independent, stateless call per report and reasoning disabled to elicit direct JSON (MiniMax forces reasoning, degraded to low effort; Gemini’s reasoning could not be disabled); (ii) the Claude Code parallel-subagent channel for Claude-Fable-5; and (iii) the ChatGPT Codex desktop-agent channel for GPT-5.5-Codex—both using platform-default sampling with file-based batch judging.

Because the two agent channels differ from the clean-API path in sampling, reasoning mode, batching and harness wrapping, we added two clean-API re-runs through the same OpenRouter channel as the other API models: Claude-Opus-4.8 (anthropic/claude-opus-4-8) and GPT-5.5 (openai/gpt-5.5), each reasoning-disabled with one stateless call per report. This yields a same-model, two-channel comparison for OpenAI (GPT-5.5-Codex vs GPT-5.5-API) that directly estimates the channel effect, and a cheap, fully reproducible Anthropic data point (Claude-Opus-4.8). Note that the current frontier Anthropic and OpenAI models do not accept a temperature parameter, so a temperature-0 setting identical to the other ten models is not attainable for these configurations by design; this is reported as a limitation. The roster thus comprises 14 model configurations (12 clean-API + 2 agent-channel). Because Claude-Fable-5 was reached through the study’s own Claude Code agent, it served as both an evaluation instrument and an evaluated system; this potential conflict was mitigated by the blinded prompt, the frozen objective gold standard, and the inclusion of the clean-API Claude-Opus-4.8 as the primary, fully reproducible Anthropic data point.

### Prompting

A single frozen prompt, built on the CRISPE framework, cast the model as a “senior nuclear-medicine physician”, supplied the six error definitions and the scoring rubric, and required a strict JSON object reporting the count of each error type and the overall score, with no explanation. The identical blinded system prompt (Supplementary Methods S3) was used across all channels. Each report was submitted in an independent session to exclude cross-report context.

### Human expert baseline

To benchmark models against human readers and to establish the agreement achievable between experts, four attending nuclear-medicine physicians—two with 10 years’ and two with 5 years’ ^18^F-FDG PET/CT reporting experience—independently applied the identical QC instrument (six binary error flags and a 1–5 overall score) to a stratified random subset of 100 reports (60 injected-error, 40 finalised), blinded to injection status. Within each seniority pair, one physician annotated the first 50 reports of the subset and the other the remaining 50 (each report thus receiving one 10-year and one 5-year rating); the two physicians’ ratings were pooled per seniority group for analysis. Because each physician rated a disjoint half, within-tier inter-rater reliability could not be assessed; the between-physician agreement reported below is therefore between the pooled 10-year and 5-year readings of each report (across the two reader pairs). For a like-for-like comparison, all models were re-scored on these same 100 reports; subset values are reported separately from the full-cohort results and are not cross-read with them.

### Statistical analysis

Data were processed in Python 3.13 with SciPy. With the injection log as reference, each report × error type was treated as a binary classification (“error present” = positive), and accuracy, precision (P), recall (R) and F1 (= 2PR/[P+R]) were computed per error type and per arm; the macro-averaged F1 is the unweighted mean of the six per-category F1 values. Agreement between each model’s overall score and the reference score was quantified by the intraclass correlation coefficient (ICC[2,1]; two-way random, single measure, absolute agreement), interpreted by the Koo & Li bands (<0.50 poor, 0.50–0.75 moderate, >0.75 good).□

To meet the inferential bar for this study, we added: (i) bootstrap 95% confidence intervals for macro-F1 and ICC for every model, by resampling reports with replacement stratified by arm (2,000 resamples, fixed seed); (ii) Spearman’s ρ between macro-F1 and ICC across models, with a bootstrap 95% CI, to test detection–calibration dissociation; (iii) a pre-specified region comparison restricted to the 12 clean-API configurations (the two agent-channel models were excluded from inference and shown only as reference points), testing the domestic-minus-international difference in macro-F1, ICC and Chinese-character-error F1 by a Monte-Carlo permutation test (10,000 label permutations), with Mann–Whitney U as a check and a bootstrap CI for the difference; (iv) a channel-effect analysis reporting the bootstrap-CI difference in macro-F1 and ICC between each agent-channel model and its clean-API counterpart; and (v) Benjamini–Hochberg FDR correction across the four pre-specified tests (region macro-F1, region ICC, region Chinese-character-error F1 and the dissociation ρ). All per-model and per-category values are reported descriptively with confidence intervals rather than multiplicity-corrected p-values. P < 0.05 was considered significant.

## Data availability

The de-identified report texts contain protected clinical information and cannot be shared publicly; the controlled-injection gold-standard log, aggregated per-model results and analysis outputs are available from the corresponding author on reasonable request, subject to institutional data-governance approval.

## Code availability

The full evaluation and analysis pipeline—de-identification, controlled error-injection sample construction, the frozen prompt, model-access configuration, scoring, statistical analysis and figure generation—is available at https://github.com/WJB1425/petqc. The repository is private during peer review (access for editors and reviewers on request) and will be made public and archived with a citable DOI (Zenodo) upon acceptance. API credentials and clinical report text are excluded from the repository; de-identified data are available from the corresponding author on reasonable request (see Data availability).

## Acknowledgements

Not applicable.

## Author contributions

J.W. and W.T. contributed equally to this work: they conceived the study, built and ran the evaluation and analysis pipeline, performed the statistical analysis and wrote the manuscript. X.M. contributed to study design and manuscript writing. H.Y. contributed to clinical data collection. Y.Y. supervised the study, provided clinical oversight, critically revised the manuscript and is the corresponding author. All authors reviewed and approved the final version.

## Ethics declarations

This retrospective study was approved by the Institutional Review Board of Shanghai Ninth People’s Hospital, Shanghai Jiao Tong University School of Medicine (approval number SH9H-2026-T390-1); the requirement for informed consent was waived because only de-identified text data were analysed.

## Reporting standards

This study is reported in accordance with the TRIPOD-LLM reporting guideline for studies using large language models^2^□; a completed TRIPOD-LLM checklist is provided as Supplementary Information, with a CLAIM (2024) checklist^2^□ as a domain-specific complement.

## Supplementary Information

- S1 model details · S2 per-arm F1 · S3 prompt · S4 statistics · TRIPOD-LLM & CLAIM checklists

## References

1. *(Nature numbered style, in citation order. Entries 1-27 carried from the validated reference set; 28-30 added for the English/benchmarking version. Re-verify all DOIs/volumes against PubMed at submission. The June-2026 proprietary-access event cited in the Discussion is a news/company statement, not a peer reviewed source, and is given as an in-text footnote rather than a numbered reference; re-check its status and wording at submission.)*

1. Tee, Q. X., Nambiar, M. & Stuckey, S. Error and cognitive bias in diagnostic radiology. J. Med. Imaging Radiat. Oncol. 66, 202–207 (2022).

2. Doo, F. X., Cook, T. S., Siegel, E. L. et al. Exploring the clinical translation of generative models like ChatGPT: promise and pitfalls in radiology, from patients to population health. J. Am. Coll. Radiol. 20, 877–885 (2023).

3. Gertz, R. J., Dratsch, T., Bunck, A. C. et al. Potential of GPT-4 for detecting errors in radiology reports: implications for reporting accuracy. Radiology 311, e232714 (2024).

4. Tian, L. P., Fei, X. L., Song, D. et al. A comparative study of large language models for intelligent quality assessment of head-and-neck CT angiography imaging reports. Chin. J. Radiol. 59, 1118–1125 (2025).

5. Koo, T. K. & Li, M. Y. A guideline of selecting and reporting intraclass correlation coefficients for reliability research. J. Chiropr. Med. 15, 155–163 (2016).

6. Doshi, R., Amin, K. S., Khosla, P. et al. Quantitative evaluation of large language models to streamline radiology report impressions: a multimodal retrospective analysis. Radiology 310, e231593 (2024).

7. Van Veen, D., Van Uden, C., Blankemeier, L. et al. Adapted large language models can outperform medical experts in clinical text summarization. Nat. Med. 30, 1134–1142 (2024).

8. Jeblick, K., Schachtner, B., Dexl, J. et al. ChatGPT makes medicine easy to swallow: an exploratory case study on simplified radiology reports. Eur. Radiol. 34, 2817–2825 (2024).

9. Adamo, S. H., Gereke, B. J., Shomstein, S. et al. From “satisfaction of search” to “subsequent search misses”: a review of multiple-target search errors across radiology and cognitive science. Cogn. Res. Princ. Implic. 6, 59 (2021).

10. Hu, B. & Zhang, L. J. Application prospects of large language models such as ChatGPT in radiology. Chin. J. Radiol. 57, 1273–1277 (2023).

11. Huang, L., Hu, J., Cai, Q. et al. The performance evaluation of artificial intelligence ERNIE bot in the Chinese national medical licensing examination. Postgrad. Med. J. 100, 952–953 (2024).

12. Salam, B., Stüwe, C., Nowak, S. et al. Large language models for error detection in radiology reports: a comparative analysis between closed-source and privacy-compliant open-source models. Eur. Radiol. 35, 4549–4557 (2025).

13. Sun, C., Teichman, K., Zhou, Y. et al. Generative large language models trained for detecting errors in radiology reports. Radiology 315, e242575 (2025).

14. Shen, H., Wu, T., Wang, F. et al. Error detection in emergency radiology reports using a large language model: multistage evaluation study. J. Med. Internet Res. 28, e86841 (2026).

15. Kim, S., et al. A multi-pass large language model framework for precise and efficient radiology report error detection. Preprint at https://arxiv.org/abs/2506.20112 (2025).

16. Fink, M. A., Bischoff, A., Fink, C. A. et al. Potential of ChatGPT and GPT-4 for data mining of free-text CT reports on lung cancer. Radiology 308, e231362 (2023).

17. Adams, L. C., Truhn, D., Busch, F. et al. Leveraging GPT-4 for post hoc transformation of free-text radiology reports into structured reporting: a multilingual feasibility study. Radiology 307, e230725 (2023).

18. Liu, J., Wang, C. & Liu, S. Utility of ChatGPT in clinical practice. J. Med. Internet Res. 25, e48568 (2023).

19. Shool, S., Adimi, S., Saboori Amleshi, R. et al. A systematic review of large language model (LLM) evaluations in clinical medicine. BMC Med. Inform. Decis. Mak. 25, 117 (2025).

20. Waisberg, E., Ong, J., Masalkhi, M. et al. GPT-4: a new era of artificial intelligence in medicine. Ir. J. Med. Sci. 192, 3197–3200 (2023).

21. Chen, L. F., Gao, X., Hou, H. T. et al. Application of generative large language models in the Chinese radiology domain. J. Front. Comput. Sci. Technol. 18, 2337–2348 (2024).

22. Li, C., Chen, Y. M., Duan, Y. et al. Evaluation of the performance of generative artificial intelligence in generating radiology reports. New Med. 55, 853–860 (2024).

23. Hu, M., Qian, J., Pan, S. et al. Advancing medical imaging with language models: featuring a spotlight on ChatGPT. Phys. Med. Biol. 69, 10TR01 (2024).

24. Lavista Ferres, J. M., Weeks, W. B., Chu, L. C., et al. Beyond chatting: the opportunities and challenges of ChatGPT in medicine and radiology. Diagn. Interv. Imaging 104, 263–264 (2023).

25. Bajaj, S., Gandhi, D. & Nayar, D. Potential applications and impact of ChatGPT in radiology. Acad. Radiol. 31, 1256–1261 (2024).

26. Temperley, H. C., O’Sullivan, N. J., Mac Curtain, B. M. et al. Current applications and future potential of ChatGPT in radiology: a systematic review. J. Med. Imaging Radiat. Oncol. 68, 257–264 (2024).

27. Sun, Y., Wang, S., Feng, S., et al. ERNIE 3.0: large-scale knowledge enhanced pre-training for language understanding and generation. Preprint at https://arxiv.org/abs/2107.02137 (2021).

28. Gallifant, J., Afshar, M., Ameen, S. et al. The TRIPOD-LLM reporting guideline for studies using large language models. Nat. Med. 31, 60–69 (2025).

29. Tejani, A. S., Klontzas, M. E., Gatti, A. A. et al. Checklist for Artificial Intelligence in Medical Imaging (CLAIM): 2024 update. Radiol. Artif. Intell. 6, e240300 (2024).

30. Mao, Y., Wang, C., Li, Y., Wang, W. & Zhang, M. Multidimensional evaluation of large language models in radiology report readability. npj Digit. Med. (2026). 10.1038/s41746-026-02589-3

31. Qin, Z., et al. Benchmarking large language models for quality control of chest radiographs and CT reports: a retrospective multimodal study. Eur. J. Radiol. 200, 112840 (2026).

32. Akinci D’Antonoli, T., Adams, L. C., Lübberstedt, J., et al. GPT-4.1 and Llama 3.3 70B fail to detect clinically relevant errors in radiology reports in zero-shot evaluation. Eur. Radiol. (2026). 10.1007/s00330-026-12697-z

