## Supplementary information for "Detection without calibration: benchmarking domestic and international large language models for quality control of Mandarin ^18^F-FDG PET/CT reports"

### Supplementary Appendix S1 — Model details, access channels and cost

This appendix details the 14 large language model configurations evaluated, their versions, how each was accessed, reasoning/sampling settings, and throughput/cost. It supports the Methods “Models and access” disclosure required for reproducibility and fair cross-model comparison. All numeric throughput/cost values derive from petqc/data/04_results/efficiency_summary.csv; public per-token prices are from petqc/config.py (international clean-API prices verified via OpenRouter on 2026-06-13). The 12 original runs were performed in June 2026; the two clean-API re-runs (Claude-Opus-4.8, GPT-5.5), added to control for access channel, were run on 2026-06-13.

#### Table S1a. Model identity, access channel, and inference settings

| # | Model (paper name) | Developer | Region | Weights | Version / model ID | Access channel | Reasoning mode | Sampling |
| --- | --- | --- | --- | --- | --- | --- | --- | --- |
| 1 | DeepSeek-V4-flash | DeepSeek | China | Proprietary API† | deepseek-chat (V4-flash, non-thinking mode) | Vendor-direct API (api.deepseek.com) | Off (non-thinking) | temp 0 |
| 2 | Qwen-Max | Alibaba (Tongyi Qianwen) | China | Proprietary API† | qwen-max | Vendor-direct API (DashScope) | Off | temp 0 |
| 3 | Hunyuan-Hy3 | Tencent | China | Proprietary API | hy3-preview | Vendor-direct API (Tencent LKEAP, /plan/v3) | Off | temp 0 |
| 4 | MiniMax-M2.5 | MiniMax | China | Proprietary API† | minimax/minimax-m2.5 | OpenRouter | Forced on → degraded to effort=low | temp 0 |
| 5 | Kimi-K2.5 | Moonshot AI | China | Proprietary API† | moonshotai/kimi-k2.5 | OpenRouter | Off | temp 0 |
| 6 | GLM-5 | Zhipu AI | China | Proprietary API† | z-ai/glm-5 | OpenRouter | Off | temp 0 |
| 7 | MiMo-V2.5-Pro | Xiaomi | China | Proprietary API† | xiaomi/mimo-v2.5-pro | OpenRouter | Off | temp 0 |
| 8 | Claude-Fable-5 | Anthropic | International | Proprietary | claude-fable-5 | Claude Code parallel subagent (self-evaluation) | effort=low (auto; not disableable) | platform default |
| 9 | GPT-5.5-Codex | OpenAI | International | Proprietary | GPT-5.5 | ChatGPT Codex desktop agent | agent default | platform default |
| 10 | Gemini-3.1-Pro | Google | International | Proprietary | google/gemini-3.1-pro-preview | OpenRouter | Could **not** be disabled (≈944 out tok/report) | temp 0 |
| 11 | Grok-4.3 | xAI | International | Proprietary | x-ai/grok-4.3 | OpenRouter | Off | temp 0 |
| 12 | Llama-4-Maverick | Meta | International | **Open weights** | meta-llama/llama-4-maverick | OpenRouter | Off | temp 0 |
| 13 | Claude-Opus-4.8 | Anthropic | International | Proprietary | anthropic/claude-opus-4.8 | OpenRouter (clean-API re-run) | Off (reasoning disabled) | platform default¶ |
| 14 | GPT-5.5 | OpenAI | International | Proprietary | openai/gpt-5.5 | OpenRouter (clean-API re-run) | Off (reasoning disabled) | platform default¶ |

† Several Chinese vendors also publish open-weight releases for some model families; the specific flagship endpoints evaluated here were accessed as hosted API services.

¶ Configurations 13–14 are clean-API re-runs of the Anthropic and OpenAI flagships through the same OpenRouter channel as configurations 4–7 and 10–12, added to control for the access-channel confound affecting configurations 8–9. The current frontier Anthropic and OpenAI models do not accept a temperature parameter (it returns an error), so a temperature-0 setting identical to the other models is not attainable for them; they were run reasoning-disabled with one independent stateless call per report.

#### Table S1b. Throughput and cost (per report and per 1,000 reports)

| Model (paper name) | Input tok/report | Output tok/report | Latency (s/report) | Cost (US$/1,000) | Public price USD (in / out, per 1M tok) |
| --- | --- | --- | --- | --- | --- |
| DeepSeek-V4-flash | 1,373 | 43.0 | 1.13 | 0.14 | 0.098 / 0.197 |
| Qwen-Max | 1,548 | 37.0 | 1.64 | 2.71 | 1.60 / 6.40 |
| Hunyuan-Hy3 | 1,305 | 37.0 | 2.51 | 0.09 | 0.063 / 0.210 |
| MiniMax-M2.5 | 1,386 | 966.0 | 23.22 | 1.32 | 0.15 / 1.15 |
| Kimi-K2.5 | 1,350 | 39.5 | 4.83 | 0.62 | 0.40 / 1.90 |
| GLM-5 | 1,407 | 45.5 | 5.13 | 0.93 | 0.60 / 1.92 |
| MiMo-V2.5-Pro | 1,548 | 38.0 | 4.46 | 0.71 | 0.435 / 0.87 |
| Claude-Fable-5 | — | — | — | 0 (Claude Code allowance) | ≈10 / ≈50‡ |
| GPT-5.5-Codex | — | — | — | 0 (ChatGPT subscription) | N/A (subscription) |
| Gemini-3.1-Pro | 1,598 | 943.9 | 59.07§ | 14.52 | 2.00 / 12.00 |
| Grok-4.3 | 1,725 | 39.0 | 1.15 | 2.25 | 1.25 / 2.50 |
| Llama-4-Maverick | 1,563 | 41.0 | 1.25 | 0.26 | 0.15 / 0.60 |
| Claude-Opus-4.8 (clean-API re-run) | 2,419 | 65.0 | 2.17 | 13.72 | 5.00 / 25.00 |
| GPT-5.5 (clean-API re-run) | 1,857 | 46.0 | 1.48 | 10.66 | 5.00 / 30.00 |

§ Gemini latency measured via the native Google API on 15 random reports (n = 13 of 15 valid, SD 33.05); its input/output tokens and cost are from the OpenRouter evaluation run, a different configuration (see Note 4).

‡ Anthropic public list price; not billed in this study (run via the Claude Code allowance).

#### Notes (methodology disclosure)

1. **Three access channels.** (i) *Clean API* — vendor-direct for DeepSeek, Qwen-Max and Hunyuan; via OpenRouter for MiniMax, Kimi, GLM-5, MiMo, Gemini, Grok and Llama — all at temperature 0 with one independent stateless call per report. (ii) *Claude Code parallel subagent* for Claude-Fable-5: each batch judged in a fresh context with platform-default sampling. (iii) *ChatGPT Codex desktop agent* for GPT-5.5: file-based batch judging with platform-default sampling. The identical blinded system prompt (Supplementary S3) and the same controlled-injection gold standard were used across all channels.
2. **Reasoning settings.** For OpenRouter calls the request asked for reasoning to be disabled to elicit direct JSON. MiniMax forces reasoning, which was therefore degraded to effort=low (hence its ≈966 output tokens/report). Gemini’s reasoning could not be disabled over OpenRouter (≈944 output tokens/report), inflating its tokens and cost roughly 20-fold versus the other API models and precluding apples-to-apples latency/output comparison. Claude-Fable-5 ran under Claude Code at effort=low (not fully disableable).
3. **Gemini access rationale.** The vendor-direct Google paid tier was throttled by a Tier-1 250-requests/day cap (only ≈91/1,000 reports completed), so the no-daily-cap OpenRouter route was used instead.
4. **Efficiency-table scope.** Claude-Fable-5 and GPT-5.5-Codex are excluded from Table S1b throughput/latency because their interactive/agent channels produce no comparable per-call token or latency logs; both incurred no cash cost (Claude Code allowance / ChatGPT subscription). International API latencies were live-timed on 15 random reports on 2026-06-13: Gemini-3.1-Pro via its **native** Google API (59.07 s/report; n = 13 of 15 valid, SD 33.05; reflecting heavy native reasoning), and Grok-4.3 (1.15 s) and Llama-4-Maverick (1.25 s) via OpenRouter. Gemini’s tokens and cost in Table S1b remain from the OpenRouter evaluation run — a different configuration from its native-API latency.
5. **Cost computation.** US$ = (input_tokens × price_in + output_tokens × price_out) / 10⁶, using each model’s public per-token USD pricing. Latency = mean over 15 timed reports per model (the seven domestic models from the original serial benchmark; the three international API models live-timed on 15 random reports, 2026-06-13).
6. **Self-evaluation safeguard.** Claude-Fable-5 is both the study’s working environment and one of the evaluated models. Bias was mitigated by blinded evaluation, an objective controlled-injection gold standard, a 1-of-14 comparative design, and freezing the prompt, gold standard and analysis code before scoring.
7. **Model selection.** Domestic models were the higher-ranked Chinese LLMs on the OpenRouter usage leaderboard (June 2026); international models comprise the four leading closed-source flagships (OpenAI, Anthropic, Google, xAI) plus an open-weights representative (Meta Llama).
8. **Clean-API re-runs and channel-effect check (configurations 13–14).** Because Claude-Fable-5 and GPT-5.5-Codex were reached through agent channels (configurations 8–9) rather than the clean API, the Anthropic and OpenAI flagships were re-run through the identical clean OpenRouter channel (Claude-Opus-4.8, GPT-5.5) on 2026-06-13, reasoning-disabled with one stateless call per report. For OpenAI this is a same-model, two-channel comparison: relative to the clean API, the Codex agent channel inflated both detection (Δmacro-F1 = +0.057, 95% CI 0.038–0.076) and calibration (ΔICC = +0.098, 95% CI 0.070–0.128). All region-level statistical inference was therefore pre-specified on the 12 clean-API configurations, with the two agent-channel configurations shown as reference points only. These re-runs also supply the Anthropic/OpenAI throughput and cost figures (Table S1b) that the agent channels could not.

### Supplementary Table S2 — Per-arm, per-category error-detection F1

*All 14 model configurations; F1 vs the controlled-injection gold standard, by arm. Pooled values are in main-text Table 1.*

**Table S2J. Junior-doctor arm — per-category F1 (n = 500).**

| Model | Descriptive | Character | Laterality | Missed dx | Logic order | Other | Macro-F1 |
| --- | --- | --- | --- | --- | --- | --- | --- |
| Claude-Opus-4.8 | 0.950 | 0.833 | 0.898 | 0.493 | 0.466 | 0.806 | 0.741 |
| Claude-Fable-5 | 0.993 | 0.777 | 0.837 | 0.283 | 0.560 | 0.879 | 0.721 |
| GPT-5.5-Codex | 1.000 | 0.599 | 0.935 | 0.601 | 0.324 | 0.755 | 0.702 |
| Gemini-3.1-Pro | 0.987 | 0.667 | 0.900 | 0.564 | 0.288 | 0.800 | 0.701 |
| GPT-5.5 | 0.874 | 0.727 | 0.881 | 0.544 | 0.298 | 0.714 | 0.673 |
| DeepSeek-V4-flash | 0.781 | 0.717 | 0.891 | 0.278 | 0.508 | 0.726 | 0.650 |
| MiMo-V2.5-Pro | 0.860 | 0.798 | 0.854 | 0.272 | 0.559 | 0.414 | 0.626 |
| GLM-5 | 0.859 | 0.809 | 0.851 | 0.352 | 0.597 | 0.281 | 0.625 |
| Hunyuan-Hy3 | 0.752 | 0.627 | 0.736 | 0.350 | 0.258 | 0.727 | 0.575 |
| MiniMax-M2.5 | 0.842 | 0.717 | 0.787 | 0.328 | 0.287 | 0.472 | 0.572 |
| Qwen-Max | 0.793 | 0.503 | 0.857 | 0.273 | 0.374 | 0.318 | 0.520 |
| Kimi-K2.5 | 0.451 | 0.629 | 0.759 | 0.348 | 0.321 | 0.570 | 0.513 |
| Llama-4-Maverick | 0.537 | 0.434 | 0.787 | 0.337 | 0.128 | 0.767 | 0.499 |
| Grok-4.3 | 0.394 | 0.411 | 0.661 | 0.292 | 0.334 | 0.642 | 0.456 |

**Table S2F. Finalised arm — per-category F1 (n = 500).**

| Model | Descriptive | Character | Laterality | Missed dx | Logic order | Other | Macro-F1 |
| --- | --- | --- | --- | --- | --- | --- | --- |
| Claude-Fable-5 | 0.867 | 0.358 | 0.947 | 0.211 | 0.144 | 0.533 | 0.510 |
| MiMo-V2.5-Pro | 0.774 | 0.634 | 0.941 | 0.174 | 0.294 | 0.167 | 0.497 |
| GLM-5 | 0.722 | 0.473 | 0.947 | 0.250 | 0.258 | 0.158 | 0.468 |
| GPT-5.5-Codex | 0.963 | 0.190 | 0.900 | 0.209 | 0.075 | 0.419 | 0.459 |
| Claude-Opus-4.8 | 0.813 | 0.500 | 0.562 | 0.298 | 0.129 | 0.381 | 0.447 |
| DeepSeek-V4-flash | 0.619 | 0.481 | 0.615 | 0.143 | 0.226 | 0.426 | 0.418 |
| Gemini-3.1-Pro | 0.929 | 0.226 | 0.643 | 0.144 | 0.062 | 0.413 | 0.403 |
| Qwen-Max | 0.634 | 0.311 | 1.000 | 0.056 | 0.154 | 0.182 | 0.390 |
| Llama-4-Maverick | 0.314 | 0.089 | 0.889 | 0.160 | 0.091 | 0.474 | 0.336 |
| MiniMax-M2.5 | 0.743 | 0.351 | 0.500 | 0.108 | 0.070 | 0.245 | 0.336 |
| GPT-5.5 | 0.456 | 0.292 | 0.720 | 0.180 | 0.065 | 0.242 | 0.326 |
| Hunyuan-Hy3 | 0.394 | 0.261 | 0.500 | 0.075 | 0.054 | 0.385 | 0.278 |
| Grok-4.3 | 0.094 | 0.086 | 0.471 | 0.042 | 0.077 | 0.333 | 0.184 |
| Kimi-K2.5 | 0.127 | 0.175 | 0.391 | 0.074 | 0.070 | 0.168 | 0.168 |

### Supplementary Methods S3 — Frozen evaluation prompt

The identical blinded system prompt below was used for every model and every report, across all access channels. It was frozen before any model was scored. The user turn supplied only the report text (Findings + Impression). Models were instructed to return a strict JSON object with the count of each of the six error types and a 1–5 overall score, with no explanation. (Original Chinese prompt; an English gloss follows for reviewers.)

#### System prompt (verbatim, Chinese)

你是一名资深核医学科医师（具10年以上PET-CT诊断经验），精通18F-FDG PET-CT全身显像报告的书写规范与质量控制。你的任务是对给定的一份PET-CT报告（含【检查所见】与【检查结论】）进行质量审核，检测其中6类错误的数目，并对报告整体质量按李克特5分量表评分。

【6类错误定义】
1. 描述错误：误用其他影像学术语描述PET-CT征象（如用"信号"或"强化"替代"糖代谢/摄取"）。
2. 书写错误：中文错别字，尤其是脏器或解剖结构名称写错（如"甲装腺"应为甲状腺，"淋巴节"应为淋巴结，"纵膈"应为纵隔，"锥体"应为椎体）。
3. 左右混淆错误：左右侧别描述前后矛盾，或检查结论中病灶侧别与检查所见不一致。
4. 诊断遗漏错误：检查所见中已明确描述的异常未在检查结论中体现。
5. 逻辑语序错误：检查结论各编号条目未遵循"颅脑→头颈→胸部→腹盆→骨骼及软组织"由头至足顺序排列。
6. 其他错误：单位错误（SUV误加单位、cm与mm误用）、标点或日期错误等。

【整体评分（李克特5分量表）】
5分：无任何常见或特殊错误，逻辑通顺；4分：无错误，仅语言稍欠通顺；3分：1~2处常见错误；
2分：3处常见错误，或存在特殊错误（左右混淆）；1分：3处以上常见错误且存在特殊错误。
（常见错误＝描述/书写/诊断遗漏/逻辑语序/其他错误；特殊错误＝左右混淆错误）

【输出要求】
严格只输出一个JSON对象，不要任何解释、前后缀或Markdown代码块。整体评分必须为1至5的整数。格式如下：
{"描述错误":0,"书写错误":0,"左右混淆":0,"诊断遗漏":0,"逻辑语序":0,"其他错误":0,"整体评分":5}

#### User turn

请审核以下PET-CT报告，输出JSON：

{report_text}

#### English gloss (for reviewers; not used at inference)

“You are a senior nuclear-medicine physician (>10 years’ PET/CT experience), expert in the writing standards and quality control of whole-body ¹⁸F-FDG PET/CT reports. Review the given report (Findings + Impression), count instances of six error types, and rate overall quality on a 1–5 Likert scale.” The six error definitions and the scoring rubric reproduce the Methods. Output is restricted to a single JSON object: {"descriptive":0,"spelling":0,"laterality":0,"missed-diagnosis":0,"logical-ordering":0,"other":0,"overall-score":5}.

#### Inference settings

Temperature 0 for all models that accept it; reasoning disabled where controllable (MiniMax forced → low effort; Gemini reasoning not disableable; the frontier Anthropic/OpenAI clean-API re-runs — Claude-Opus-4.8, GPT-5.5 — do not accept a temperature parameter and were run reasoning-disabled with default sampling). One independent, stateless call per report on the clean-API channel; the two agent-channel models (Claude-Fable-5, GPT-5.5-Codex) used platform-default sampling with file-based batch judging. max_tokens = 4000.

### Supplementary Table S4 — Inferential statistics

*Bootstrap 95% CIs (2,000 resamples, arm-stratified); region permutation test (10,000) pre-specified on the 12 clean-API configurations; channel-effect bootstrap-CI deltas; Benjamini–Hochberg FDR over the hypothesis family. Generated by analyze_stats.py (fixed seed).*

#### S4a. Bootstrap 95% CIs (macro-F1, ICC, Chinese-character-error F1)

| model | model_key | region | channel | macroF1 | macroF1_lo | macroF1_hi | ICC | ICC_lo | ICC_hi | charErrorF1 | charErrorF1_lo | charErrorF1_hi |
| --- | --- | --- | --- | --- | --- | --- | --- | --- | --- | --- | --- | --- |
| Claude-Opus-4.8 | claude_opus_api | intl | clean-API | 0.667 | 0.639 | 0.692 | 0.491 | 0.446 | 0.536 | 0.764 | 0.699 | 0.822 |
| Claude-Fable-5 | claude | intl | agent | 0.66 | 0.63 | 0.687 | 0.559 | 0.513 | 0.607 | 0.667 | 0.599 | 0.732 |
| GPT-5.5-Codex | gpt_web | intl | agent | 0.624 | 0.603 | 0.643 | 0.329 | 0.294 | 0.364 | 0.457 | 0.395 | 0.518 |
| Gemini-3.1-Pro | gemini | intl | clean-API | 0.609 | 0.586 | 0.632 | 0.244 | 0.212 | 0.276 | 0.52 | 0.459 | 0.581 |
| MiMo-V2.5-Pro | mimo | cn | clean-API | 0.602 | 0.566 | 0.633 | 0.627 | 0.573 | 0.674 | 0.766 | 0.697 | 0.824 |
| DeepSeek-V4-flash | deepseek | cn | clean-API | 0.594 | 0.558 | 0.626 | 0.609 | 0.555 | 0.656 | 0.661 | 0.589 | 0.73 |
| GLM-5 | glm | cn | clean-API | 0.586 | 0.554 | 0.615 | 0.612 | 0.562 | 0.657 | 0.733 | 0.669 | 0.79 |
| GPT-5.5 | gpt55_api | intl | clean-API | 0.567 | 0.541 | 0.592 | 0.231 | 0.198 | 0.263 | 0.597 | 0.533 | 0.658 |
| MiniMax-M2.5 | minimax | cn | clean-API | 0.507 | 0.478 | 0.532 | 0.414 | 0.359 | 0.469 | 0.631 | 0.56 | 0.697 |
| Hunyuan-Hy3 | hunyuan | cn | clean-API | 0.488 | 0.462 | 0.513 | 0.369 | 0.322 | 0.415 | 0.518 | 0.451 | 0.582 |
| Qwen-Max | qwen | cn | clean-API | 0.478 | 0.447 | 0.505 | 0.352 | 0.306 | 0.394 | 0.459 | 0.372 | 0.541 |
| Llama-4-Maverick | llama | intl | clean-API | 0.461 | 0.426 | 0.493 | 0.544 | 0.484 | 0.592 | 0.341 | 0.275 | 0.405 |
| Kimi-K2.5 | kimi | cn | clean-API | 0.391 | 0.369 | 0.415 | 0.262 | 0.224 | 0.302 | 0.481 | 0.415 | 0.547 |
| Grok-4.3 | grok | intl | clean-API | 0.356 | 0.332 | 0.379 | 0.099 | 0.078 | 0.119 | 0.265 | 0.223 | 0.309 |

#### S4b. Domestic vs international (clean-API set)

| metric | domestic_n | domestic_median | domestic_min | domestic_max | intl_n | intl_median | intl_min | intl_max | delta_cn_minus_intl | delta_lo | delta_hi | perm_P | mannwhitney_P |
| --- | --- | --- | --- | --- | --- | --- | --- | --- | --- | --- | --- | --- | --- |
| Detection macro-F1 | 7 | 0.507 | 0.391 | 0.602 | 5 | 0.567 | 0.356 | 0.667 | -0.011 | -0.024 | 0.001 | 0.8372 | 0.8763 |
| Calibration ICC(2,1) | 7 | 0.414 | 0.262 | 0.627 | 5 | 0.244 | 0.099 | 0.544 | 0.142 | 0.12 | 0.164 | 0.1726 | 0.149 |
| Chinese-character-error F1 | 7 | 0.631 | 0.459 | 0.766 | 5 | 0.52 | 0.265 | 0.764 | 0.109 | 0.081 | 0.136 | 0.263 | 0.4318 |

#### S4c. Channel effect (agent vs clean API)

| comparison | agent_model | api_model | macroF1_agent | macroF1_api | macroF1_delta | macroF1_delta_lo | macroF1_delta_hi | ICC_agent | ICC_api | ICC_delta | ICC_delta_lo | ICC_delta_hi |
| --- | --- | --- | --- | --- | --- | --- | --- | --- | --- | --- | --- | --- |
| GPT-5.5: agent (Codex) vs clean API (same model, pure channel effect) | GPT-5.5-Codex | GPT-5.5(API) | 0.624 | 0.567 | 0.057 | 0.038 | 0.076 | 0.329 | 0.231 | 0.098 | 0.07 | 0.128 |
| Anthropic: Fable-5 agent vs Opus-4.8 clean API (channel + model) | Claude-Fable-5 | Claude-Opus-4.8(API) | 0.66 | 0.667 | -0.007 | -0.034 | 0.018 | 0.559 | 0.491 | 0.069 | 0.019 | 0.117 |

#### S4d. Detection-calibration association (Spearman) and BH-FDR family

| set | n_models | spearman_rho | rho_lo | rho_hi | p_value |
| --- | --- | --- | --- | --- | --- |
| all_models | 14 | 0.38 | 0.297 | 0.547 | 0.1799 |

| hypothesis | raw_P | rank | BH_FDR |
| --- | --- | --- | --- |
| Region: calibration ICC (clean-API set) | 0.1726 | 1 | 0.6903 |
| Detection-calibration dissociation: Spearman rho != 0 | 0.1799 | 2 | 0.3598 |
| Region: Chinese-character-error F1 (clean-API set) | 0.263 | 3 | 0.3506 |
| Region: detection macro-F1 (clean-API set) | 0.8372 | 4 | 0.8372 |

### TRIPOD-LLM reporting checklist (completed)

Reporting follows the TRIPOD-LLM guideline (Gallifant et al., *Nat. Med.* 2025). This is an **evaluation** study of off-the-shelf LLMs (no model development/fine-tuning); items specific to model development are marked N/A. Section references are to the main manuscript and Supplementary Information (S1 = model details; S3 = prompt).

| # | TRIPOD-LLM item | Addressed | Location |
| --- | --- | --- | --- |
| 1 | Title identifies the study as evaluating LLM(s) for a clinical task | Yes | Title |
| 2 | Abstract: objective, data, models, metrics, key results, conclusion | Yes | Abstract |
| 3 | Background and rationale; intended use and users | Yes | Introduction |
| 4 | Objectives / hypotheses (detection, calibration, region, channel) | Yes | Introduction (final paragraph) |
| 5 | Data source, setting, eligibility, de-identification | Yes | Methods — Report source and de-identification |
| 6 | Task definition and reference standard (controlled error injection) | Yes | Methods — Error taxonomy; Controlled error injection |
| 7 | Sample size / data partitioning (1,794 → 1,000; two 500-report arms) | Yes | Methods — Sampling and two-arm design |
| 8 | **LLM identity and version** (name, snapshot, developer, weights) | Yes | Methods — Models and access; Table S1a |
| 9 | **Access and query details** (channel, API/endpoint, access dates) | Yes | Methods — Models and access; Table S1a; S1 Note 8 |
| 10 | **Prompt(s) and prompt-engineering strategy** (verbatim, frozen, zero-shot) | Yes | Methods — Prompting; Supplementary Methods S3 |
| 11 | **Inference settings** (temperature, reasoning mode, stateless calls, max tokens) | Yes | Methods — Models and access; S3; S1 Notes 2, 8 |
| 12 | Output handling / parsing (strict JSON; coercion rules) | Yes | Methods — Prompting; Code availability |
| 13 | Performance metrics defined (F1, ICC(2,1), κ) with rationale | Yes | Methods — Statistical analysis |
| 14 | Uncertainty quantification (bootstrap 95% CIs) | Yes | Methods — Statistical analysis; Tables 1–2 |
| 15 | Comparator / human baseline (four attending physicians, two per seniority tier) | Yes | Methods — Human expert baseline; Results — Human expert baseline |
| 16 | Statistical comparison methods (permutation, channel-effect, multiplicity) | Yes | Methods — Statistical analysis; Results — Domestic vs international |
| 17 | Participant/data flow and characteristics | Yes | Fig. 1; Methods |
| 18 | Results: per-model and per-category performance with CIs | Yes | Results; Tables 1–5; Figs 2–3 |
| 19 | Limitations, generalisability, intended deployment, ethics | Yes | Discussion (limitations); Ethics declarations |
| — | **Open science**: code/data availability; reproducibility (fixed seeds) | Yes | Methods — Data/Code availability; fixed seed 20260603 |
| — | **Human oversight / intended workflow** (human-in-the-loop cascade) | Yes | Discussion (deployment); Conclusion |
| — | Patient/public involvement | N/A | Retrospective de-identified text; none |
| — | Model development / fine-tuning items | N/A | Off-the-shelf evaluation; zero-shot |
| — | Funding and conflicts of interest | Yes | End matter |

**Self-evaluation disclosure (study-specific).** One evaluated configuration (Claude-Fable-5) was reached through the same Claude Code environment used to build the pipeline. Bias mitigation: blinded evaluation, an objective controlled-injection gold standard not defined by any evaluated model, a 1-of-14 comparative design, frozen prompt/gold/analysis code, and a clean-API re-run of the Anthropic flagship (Claude-Opus-4.8) through the standard channel (S1 Note 8).

*A CLAIM (2024) checklist is additionally provided as a domain-specific (radiology/imaging) complement.*

### CLAIM (2024) checklist — Checklist for Artificial Intelligence in Medical Imaging

Completed against the CLAIM 2024 update (Mongan, Kohli, Houston et al., *Radiol. Artif. Intell.* 2024) as a domain-specific (radiology/imaging-AI) complement to the primary TRIPOD-LLM checklist. This study evaluates off-the-shelf LLMs on the **text** of ¹⁸F-FDG PET/CT reports; it performs no model training and no pixel-level image analysis, so imaging-acquisition, segmentation and training items are marked **N/A** with rationale. Section references are to the main manuscript (M), Supplementary Information (S1 = model details, S3 = prompt) and the code repository.

#### Title / Abstract

| # | Item | Status | Location |
| --- | --- | --- | --- |
| 1 | Identifies study as AI applied to a clinical imaging-report task | Yes | Title |
| 2 | Structured/abstract summary of design, methods, results, conclusions | Yes | Abstract |

#### Introduction

| # | Item | Status | Location |
| --- | --- | --- | --- |
| 3 | Scientific and clinical background; rationale | Yes | Introduction |
| 4 | Study objectives and hypotheses | Yes | Introduction (final paragraph) |

#### Methods — study design & data

| # | Item | Status | Location |
| --- | --- | --- | --- |
| 5 | Prospective/retrospective; study goal (evaluation/benchmark) | Yes | Methods — retrospective evaluation |
| 6 | Data source(s) | Yes | Methods — Report source |
| 7 | Eligibility / inclusion–exclusion criteria | Yes | Methods — Report source (exclusions); Sampling |
| 8 | Data pre-processing steps | Yes | Methods — de-identification; Findings+Impression only |
| 9 | Selection of data subsets | Yes | Methods — Sampling and two-arm design |
| 10 | Definitions of data elements (six error types, score) | Yes | Methods — Error taxonomy; S3 |
| 11 | De-identification methods | Yes | Methods — Report source and de-identification |
| 12 | Handling of missing data | Yes | Methods — Statistical analysis (unscored excluded); code |
| 13 | **Imaging protocol / acquisition parameters** | **N/A** | Report **text** only; no image data analysed |

#### Methods — ground truth (reference standard)

| # | Item | Status | Location |
| --- | --- | --- | --- |
| 14 | Definition of reference standard | Yes | Methods — Controlled error injection (injection log = gold) |
| 15 | Rationale for choosing the reference standard | Yes | Methods — Controlled error injection; Discussion (limitations) |
| 16 | Source of ground-truth annotations | Yes | Methods — Controlled error injection (programmatic, logged) |
| 17 | Annotation tools / process | Yes | Methods; Code availability (build_eval_set.py) |
| 18 | Inter-rater variability (where human annotation used) | Yes | Methods — Human baseline; Results — Human expert baseline (between-physician ICC, κ) |

#### Methods — data partitions

| # | Item | Status | Location |
| --- | --- | --- | --- |
| 19 | How data were assigned to partitions/arms | Yes | Methods — Sampling (disjoint 500 + 500, seed 20260603) |
| 20 | Level at which partitions are disjoint (report level) | Yes | Methods — Sampling |
| 21 | Use of a held-out / independent test set | Yes (subset) | Methods — Human baseline (100-report subset) |

#### Methods — model

| # | Item | Status | Location |
| --- | --- | --- | --- |
| 22 | Detailed model description (names, versions, weights) | Yes | Methods — Models and access; Table S1a |
| 23 | Software libraries, frameworks, packages | Yes | Methods (Python 3.13, SciPy); Code availability |
| 24 | Initialisation / model access & query details | Yes | Methods — Models and access; S1 Notes 1–2, 8 |
| 25 | Hardware / inference settings (temp, reasoning, max tokens) | Yes | Methods; S3; S1 |
| 26–29 | **Model training, hyper-parameters, training data, regularisation** | **N/A** | Off-the-shelf, zero-shot; no training/fine-tuning |

#### Methods — evaluation & statistics

| # | Item | Status | Location |
| --- | --- | --- | --- |
| 30 | Metrics of model performance | Yes | Methods — Statistical analysis (F1, ICC, κ) |
| 31 | Statistical measures of significance & uncertainty | Yes | Methods (bootstrap CIs, permutation, channel-effect, BH-FDR) |
| 32 | Robustness / sensitivity analysis | Yes | Results — Domestic vs international — channel-effect re-run; per-arm analysis |
| 33 | Methods for explainability / interpretability | Partial | Per-category and over-flagging analyses |
| 34 | Validation or testing on external data | No | Single-centre; external validation stated as future work (Discussion) |

#### Results

| # | Item | Status | Location |
| --- | --- | --- | --- |
| 35 | Flow of data through the study | Yes | Fig. 1; Methods |
| 36 | Demographic / clinical characteristics of the data | Partial | Methods (single-centre, head-and-neck-weighted cohort) |
| 37 | Model performance with uncertainty intervals | Yes | Results; Tables 1–5; Figs 2–3 |
| 38 | Failure / error analysis | Yes | Error-type patterns; over-flagging and harsh scoring |

#### Discussion

| # | Item | Status | Location |
| --- | --- | --- | --- |
| 39 | Study limitations including bias/generalisability | Yes | Discussion (limitations paragraph) |
| 40 | Implications for practice; intended clinical workflow | Yes | Discussion (deployment); Conclusion |

#### Other information

| # | Item | Status | Location |
| --- | --- | --- | --- |
| 41 | Registration / protocol | N/A | Methodological benchmark; no prospective registration |
| 42 | Source of funding and role of funders; conflicts | Yes | End matter (Funding; Competing interests) |
| — | Data and code availability | Yes | Methods — Data/Code availability |

**Domain note.** Items concerning image acquisition, reconstruction, segmentation and pixel-level annotation (CLAIM’s imaging-specific core) are not applicable: the model input is the de-identified free text of the Findings and Impression sections, and the task is textual quality control rather than image interpretation. CLAIM is provided as a complementary checklist because the clinical domain is radiology/nuclear medicine; the primary reporting guideline for this LLM-evaluation study is TRIPOD-LLM (see TRIPOD_LLM_checklist.md).
